# A Chinese host genetic study discovered type I interferons and causality of cholesterol levels and WBC counts on COVID-19 severity

**DOI:** 10.1101/2021.06.04.21258335

**Authors:** Huanhuan Zhu, Fang Zheng, Linxuan Li, Yan Jin, Yuxue Luo, Zhen Li, JingYu Zeng, Ling Tang, Zilong Li, Ningyu Xia, Panhong Liu, Dan Han, Ying Shan, Xiaoying Zhu, Siyang Liu, Rong Xie, Yilin Chen, Wen Liu, Longqi Liu, Xun Xu, Jian Wang, Huanming Yang, Xia Shen, Xin Jin, Fanjun Cheng

**Author notes:** These authors are co-corresponding authors.Correspondence: Xin Jin or Fanjun Cheng. These authors contributed equally to this work (Huanhuan Zhu, Fang Zheng, Linxuan Li, Yan Jin, Yuxue Luo, Zhen Li).

## Abstract

As of early May 2021, the ongoing pandemic COVID-19 has caused over 160 million of infections and over 3 million deaths worldwide. Many risk factors, such as age, gender, and comorbidities, have been studied to explain the variable symptoms of infected patients. However, these effects may not fully account for the diversity in disease severity. Here, we present a comprehensive analysis of a broad range of patients’ laboratory and clinical assessments to investigate the genetic contributions to COVID-19 severity. By performing GWAS analysis, we discovered several concrete associations for laboratory features. Based on these findings, we performed Mendelian randomization (MR) analysis to investigate the causality of laboratory traits on disease severity. From the MR study, we identified two causal traits, cholesterol levels and WBC counts. The functional gene related to cholesterol levels is *ApoE* and people with particular *ApoE* genotype are more likely to have higher cholesterol levels, facilitating the process that SARS-CoV-2 binds on its receptor ACE2 and aggravating COVID-19 disease. The functional gene related to WBC counts is *MHC* system that plays a central role in the immune system. The host immune response to the SARS-CoV-2 infection greatly affects the patients’ severity status and clinical outcome. Additionally, our gene-based and GSEA analysis revealed interferon pathways, including type I interferon receptor binding, regulation of IFNA signaling, and SARS coronavirus and innate immunity. We hope that our work will make a contribution in studying the genetic mechanisms of disease illness and serve as useful reference for the clinical diagnosis and treatment of COVID-19.

## Introduction

The coronavirus disease 2019 (COVID-19) is a contagious disease caused by severe acute respiratory syndrome coronavirus 2 (SARS-CoV-2). Since the late December of 2019, the COVID-19 has spread rapidly around the world leading to an ongoing pandemic. As of early May 2021, reported to the world health organization (WHO), there have been over 160 million confirmed cases of COVID-19, including over 3 million deaths. Common symptoms include fever, cough, and fatigue. Meanwhile, the symptoms could be largely variable; for example, about a third of patients do not develop noticeable symptoms; of patients who develop noticeable symptoms, 81% develop mild to moderate symptoms, while 14% develop severe symptoms, and 5% suffer critical symptoms ^1^. Many key factors have been reported to be associated with COVID-19 severity, such as age, sex, and comorbidities. Specifically, older people are more likely to be infected by SARS-CoV-2 and experience more severe symptoms. Global data indicate higher COVID-19 fatality rates among men than women. Most countries reported that the male case fatality is more than 1.0 higher than that of female ^2,3^. Besides, comorbidities have a critical role in poor outcomes, severity of disease and high fatality rate of COVID-19 cases ^4^. However, these risk factors cannot fully explain the clinical variability among the patients.

Many recent studies turn their attention to the host genetic backgrounds and believe that the genetic factor may play an essential role in determining the host responses to SARS-CoV-2 ^5-9^. By performing large-scale genome-wide association studies (GWAS) of COVID-19 clinical phenotypes, several disease-associated variants and genes were identified and summarized by the Host Genetics Initiative (HGI) ^9^, such as the rs11385942 (*SLC6A20*), rs657152 (*ABO*), and rs2236757 (*IFNAR2*) ^10,11^. Among these findings, *SLC6A20* encodes a proline transporter and is functionally associated with *ACE2* (angiotensin-converting enzyme 2), which encodes the well-known SARS-CoV-2 receptor ^10^; gene *IFNAR2* encodes one type of I interferons that is essential to the establishment of antiviral state and intensifying to antiviral response^12^. However, most of existing GWAS studies are based on the European populations, or meta-analysis with multiple populations. It is a pity that the genomic studies based on Asian populations, especially Chinese population, are relatively few. Wang et al. (2020) reported the first host genetic study in the Chinese population of 332 COVID-19 patients and suggested some relatively significant genetic loci as candidate variants associated with severity status ^5^. However, their study did not identify any genome-wide significant genetic variants (p-values < 5E-08) due to a small sample size and limited effects of single variants.

In our study, we performed extensive GWAS analyses for a wide range of laboratory assessments measured from the blood test of 466 COVID-19 patients. Even with a relatively small sample size, our GWAS results indeed identified several concrete genome-wide significant associations, which are either the first replication study for a previously reported signal or the first discovery in Chinese population based on genetic analysis. These variant-trait associations include rs1801020 (*F12*) with activated partial thromboplastin time, rs56393506 (*LPA*) with lipoprotein-A, rs28946889 (*UGT1A* complex) with total/indirect bilirubin levels, rs7412 (*ApoE*) with low-density lipoprotein cholesterol levels (LDL-C), and rs9268517 (*BTNL2*) with white blood cell counts (WBC). Based on these substantial findings, we implemented Mendelian randomization (MR) to examine whether these traits are causal factors to the COVID-19 susceptibility and severity.

MR uses genetic variants as instrumental variables to determine whether an observational association between a risk exposure and an outcome disease is also a causality ^13^. In recent years, MR has rapidly gained popularity in epidemiology and medical research, because of the ever-expanding genetic databases and well-powered GWAS studies on a large number of traits. We selected the above traits with established associations as exposure variables and the COVID-19 severity status released by the HGI database as the outcome variable. By performing MR analysis, we uncovered the causal associations of LDL-C and WBC on the disease severity. The cholesterol level and WBC counts-related genes are *ApoE* and *MHC* (major histocompatibility complex) system, respectively. We further studied the genetic architecture of how *ApoE* affects the severe illness of COVID-19 and revealed that *ApoE* could affect the severity of COVID-19 by influencing cholesterol levels in the peripheral tissues. Specifically, people with particular *ApoE* genotype are more likely to have higher levels of cholesterol, which leads the plasma membrane to form more lipid rafts. Compared to other people, these people are more vulnerable to SARS-CoV-2 as more lipid rafts would facilitate the binding of the virus to its target receptor ACE2 ^14^. The genetic mechanisms of how *MHC* system contributes to the susceptibility and severity of COVID-19 are mainly through activating and regulating the immune system. Specifically, MHC family works as an immune activator and directly triggers the proliferation of lymphocyte cells. As the number of lymphocyte cell increases, immune system springs into action against SARS-CoV-2. Therefore, the abnormal change of immune cell counts and the MHC expression levels often used as indicators of the severity of COVID-19 symptom ^15^.

We further performed gene-based studies and gene-set enrichment analysis (GSEA) based on the GWAS single-variant associations for testing COVID-19 severity. As a result, we successfully discovered four functional pathways: regulation of IFNA signaling, SARS coronavirus and innate immunity, type I interferon receptor binding, and overview of interferons-mediated signaling pathway. The type I interferon (IFN-I) can bind to the receptor on the surface of immune cell membrane, mobilize and enhance the activity of immune cells, prevent the transmission of virus between cells, and clear the cells infected by the virus ^16,17^. A study reported that the characteristic of severe COVID-19 cases was the IFN-I response and the mouse model of SARS-CoV-2 infection showed that the timing of IFN-I response is the key factor to determine the outcome of infection ^18^. In March 2020, the National Health Commission and the National Administration of Traditional Chinese Medicine issued the COVID-19 diagnosis and treatment plan, IFN-I is one of the main antiviral drugs ^19^. To the best of our knowledge, this is the first time that the interferons-related pathways are uncovered from the genetic studies of COVID-19 patients in Chinese population.

In summary, we succeeded in identifying genome-wide significant associations between genetic variants and laboratory traits measured from Chinese ancestry COVID-19 patients. Most of these findings were supported by previous literatures. On the basis of these concrete associations, we conducted MR analysis and detected two candidate genes, *ApoE* and *MHC* system, which influence the severity status by acting on cholesterol levels and WBC counts, respectively. Besides, we identified four interferons functional pathways that directly determine the COVID-19 disease severity based on genomic studies of infected patients. These findings provide new insights in studying the genetic mechanisms of COVID-19 susceptibility and severity. We hope that our work will serve as useful reference for academic field and make contribution to investigate the COVID-19 disease and finally stop the pandemic.

## Results

### Basic information of the enrolled patients

After quality control (Materials and Methods), there were 466 patients used for analysis, of which 229 were males (49.1%) and 237 were females (50.9%) (Figure 1A). The age of patients ranged from 23 to 97, composing with 20-39 (8.5%), 40-59 (31.1%), 60-79 (51.1%), and 80-99 (9.2%) (Figure 1A). According to the patients’ severity of illness at the time of admission to the hospital, they were classified into four categories as mild (*N* = 6, 1.29%), moderate (*N* = 164, 35.19%), severe (*N* = 227, 48.71%), and critical (*N* = 69, 14.81%). The method of classifying severity followed the criteria made by the National Health Commission of the People’s Republic of China ^20^. We further broadly defined the mild group as mild and moderate patients (*N* = 170) and the severe group as severe and critical patients (*N* = 296) (Figures 1B-1C). We then fitted a single factor linear regression model and statistically proved that age was a risk factor for severe symptoms of COVID-19 (z-score = 4.146, p-value = 3.38E-05). Besides, we performed a Fisher’s exact test to test the independence of patients’ gender and severity and found a significant correlation (OR = 1.59, p value = 0.016), revealing a higher propensity for severe in males with COVID-19. These phenomena were consistent with reports from previous literatures ^21,22^.

**Figure 1.**
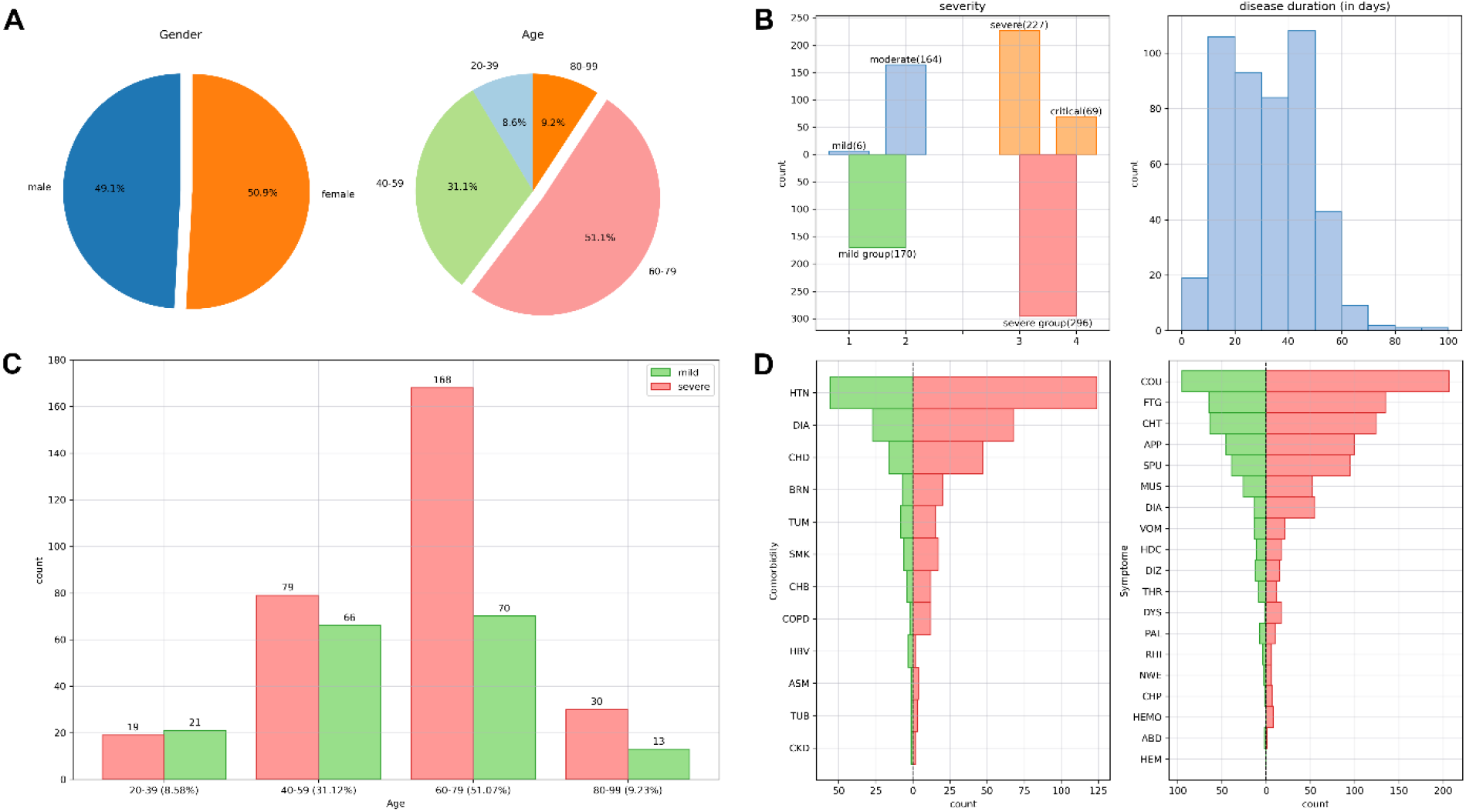
Basic clinical information of COVID-19 patients *Notes*. (A) Pie diagrams for sex ratio and age distribution of 466 samples. (B) Bar charts for severity category and histogram of hospitalized days. In severity chart, the blue and green bars indicate the mild group, orange and red bars indicate the severe group. (C) Bar chart for the counts of severity in each age range. (D) Bar charts for the distributions of comorbidities and symptoms. For the comorbidities, HTN, CHD, BRN, TUM, SMK, CHB, COPD, HBV, ASM, TUB, and CKD indicate hypertension, coronary heart disease, brain infarction, tumor, smoking history, chronic bronchitis, chronic obstructive pulmonary disease, Hepatitis B virus, asthma, tuberculosis, and chronic kidney disease, respectively. For the symptoms, COU, FTG, CHT, APP, SPU, MUS, DIA, VOM, HDC, DIZ, THR, DYS, PAL, RHI, NEW, CHP, HEMO, ABD, and HEMO indicate cough, fatigue, chest tightness, poor appetite, sputum, muscle ache, diarrhea, vomiting, headache, dizziness, sore throat, dyspnea, palpitation, rhinorrhea, night sweating, chest pain, hemoptysis, abdominal pain, and hematemesis, respectively.

More than 50% of the patients (*N* = 288) had at least one comorbidity prior to admission to the hospital, and the most frequent ones were hypertension (*N* = 180, 38.63%), diabetes (*N* = 95, 20.38%), and coronary heart disease (*N* = 63, 13.52%). The distribution of comorbidities among mild and severe patients is provided in Figure 1D. We then tested whether the presence or absence of comorbidities would affect the patients’ severity by performing a Fisher’s exact test and found that having comorbidities is a risk determinant to develop severe symptoms (OR = 1.86, p = 2.09E-03). This conclusion has been supported by many studies ^23,24^. Most of the patients experienced various COVID-19 symptoms, including cough (*N* = 302, 64.81%), fatigue (*N* = 200, 42.92%), and chest tightness (*N* = 188, 40.34%). We also reported the distribution of symptoms among mild and severe patients (Figure 1D).

### Genome-wide association analysis of laboratory features

We first evaluated the imputation accuracy of genetic variants by two measurements: imputation score and correlation with chip array sequencing. After quality control (Materials and Methods), a total of 6,349,370 variants were selected for further analysis and 99.6% of these variants had imputation score over 0.8 based on reference panel as EAS population from the 1000 Genome Project. In addition, 214 patients were sequenced with high depth and high coverage. We took overlap of variants between their chip array genotypes and imputed genotypes and it yield 479,823 sites. Over 98.1% patients had correlation coefficients above 0.8 across these genetic sites. With a mean sequencing depth of 17.8x, we finally tested a total of 6,185,321 autosomal variants and 164,049 X-chromosome variants for association with 78 quantitative laboratory traits in 466 COVID-19 patients. These laboratory measurements were grouped into 10 distinct categories (Table 1): hematological (n = 22), anticoagulation (n = 7), electrolyte (n = 7), lipid (n = 7), protein (n = 4), liver-related (n = 12), kidney-related (n = 3), heart-related (n = 8), inflammation (n = 3), and other biochemical (n = 5). The study workflow is designed as in Figure 2. When we applied multiple-testing correction to the number of the studied traits, 5 variant-trait associations were significant signals (p-value < 5E-08/78 = 6.41E-10), 4 out of which were previously identified in either European, Asian, or both populations (Table 2). These associations include rs1801020 (*F12*, p-value = 4.13E-16) with activated partial thromboplastin time (APTT), rs56393506 (*LPA*, p-value = 1.97E-14) with lipoprotein-A (LpA), rs28946889 (*UGT1A* complex, p-value = 5.08E-14) with total bilirubin levels (Tbil), and rs28946889 (*UGT1A* complex, p-value = 1.51E-16) with indirect bilirubin levels (Ibil). The Manhattan plots and QQ-plots were drawn for APTT, LpA, and Ibil with the CMplot package in R ^25^ and provided in Figure 3. A novel association was rs11032789 (*EHF*, p-value = 6.40E-10) with apoprotein A (apoA). Even though the association between rs7412 (*ApoE*, pvalue = 2.30E-08) and low-density lipoprotein cholesterol levels (LDL-C) did not reach the study-wide significance threshold, it had been widely identified in European, Asian, and Chinese populations. The association between rs9268517 (*BTNL2*, p-value = 4.05E-08) with white blood cell counts (WBC) did not pass the threshold either. Considering that *BTNL2* encodes a major histocompatibility complex (MHC) class II protein and is involved in immune surveillance, we considered this identified association was a worth-investigating signal. The Manhattan plots for LDL-C and WBC were provided in Figure 4A and Figure 5A, respectively.

**Table 1.**
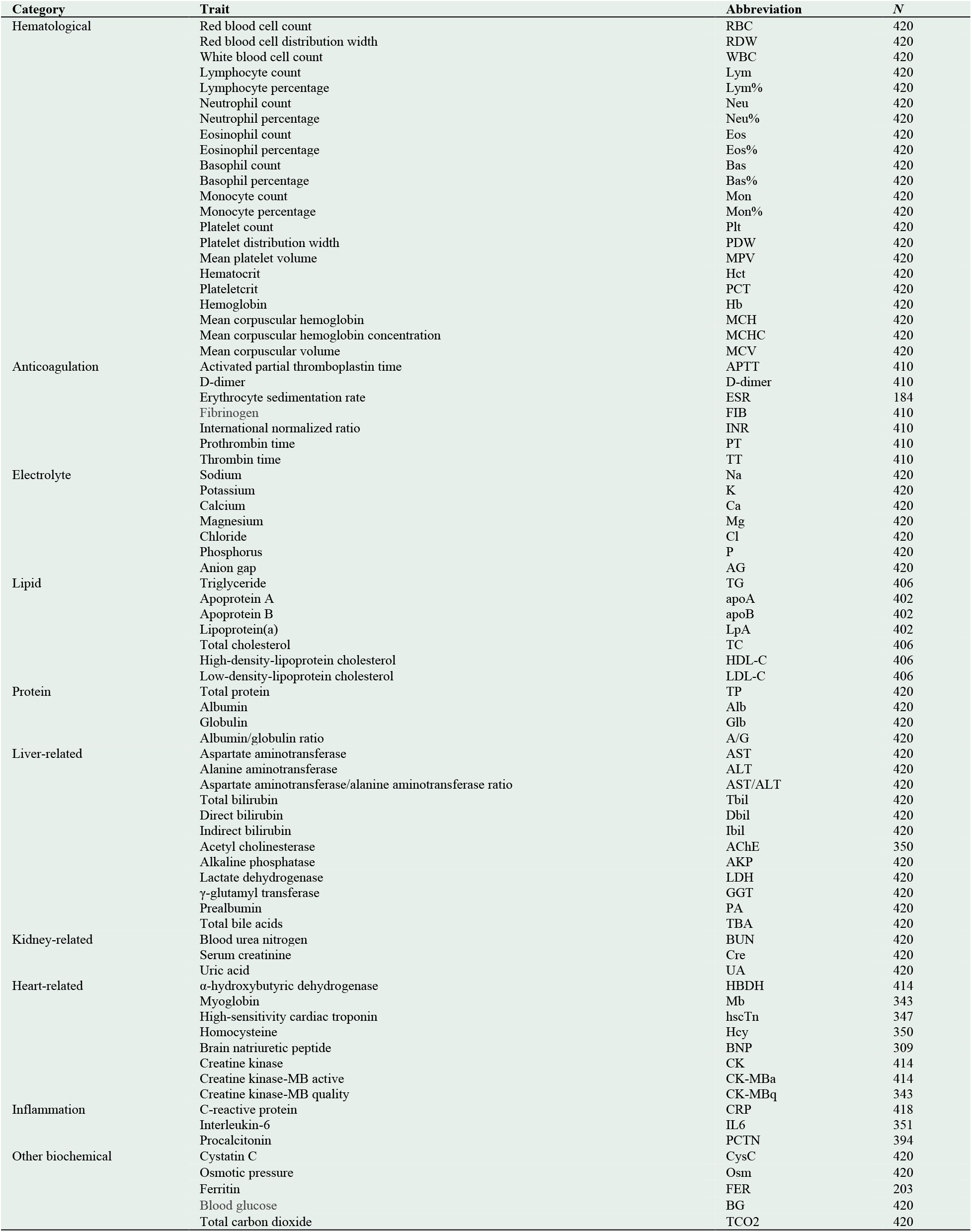
Overview of the tested laboratory assessments

**Table 2.** The concrete associations identified from single-variant GWAS analysis

| Trait | SNP | CHR | POS | REF | ALT | Mapped/Closest<br>Gene | AF | R2 | beta | se | p-value | N | European | Asian | Chinese |
| --- | --- | --- | --- | --- | --- | --- | --- | --- | --- | --- | --- | --- | --- | --- | --- |
| APTT | rs1801020 | 5 | 177409531 | A | G | *F12* | 0.254 | 1.000 | -0.606 | 0.071 | 4.13E-16 | 410 | X | √ | X |
| LpA | rs56393506 | 6 | 160668275 | C | T | *LPA* | 0.114 | 0.998 | 0.817 | 0.103 | 1.97E-14 | 402 | √ | X | X |
| Tbil | rs28946889 | 2 | 233762816 | G | T | *UGT1A* Complex | 0.400 | 0.996 | -0.521 | 0.067 | 5.08E-14 | 420 | X | √ | X |
| Ibil | rs28946889 | 2 | 233762816 | G | T | *UGT1A* Complex | 0.400 | 0.996 | -0.585 | 0.068 | 1.51E-16 | 420 | X | √ | X |
| apoA | rs11032789 | 11 | 34624907 | T | G | *EHF* | 0.040 | 1.000 | 0.960 | 0.151 | 6.40E-10 | 402 | X | X | X |
| LDL-C | rs7412 | 19 | 44908822 | C | T | *ApoE* | 0.092 | 0.998 | -0.652 | 0.114 | 2.30E-08 | 406 | √ | √ | √ |
| WBC | rs9268517 | 6 | 32411963 | C | T | *BTNL2, HLA-DRA* | 0.067 | 1.000 | 0.721 | 0.129 | 4.05E-08 | 420 | X | X | X |

*Notes.* AF indicates the allele frequency for the effect/alternate allele; R2 indicates the imputation score based on EAS population from the 1000 Genome Project; N is the sample size used in GWAS analysis; “√” and “X” indicate the corresponding associations were previously reported and not reported in a population based on genomic studies, respectively.

**Figure 2.**
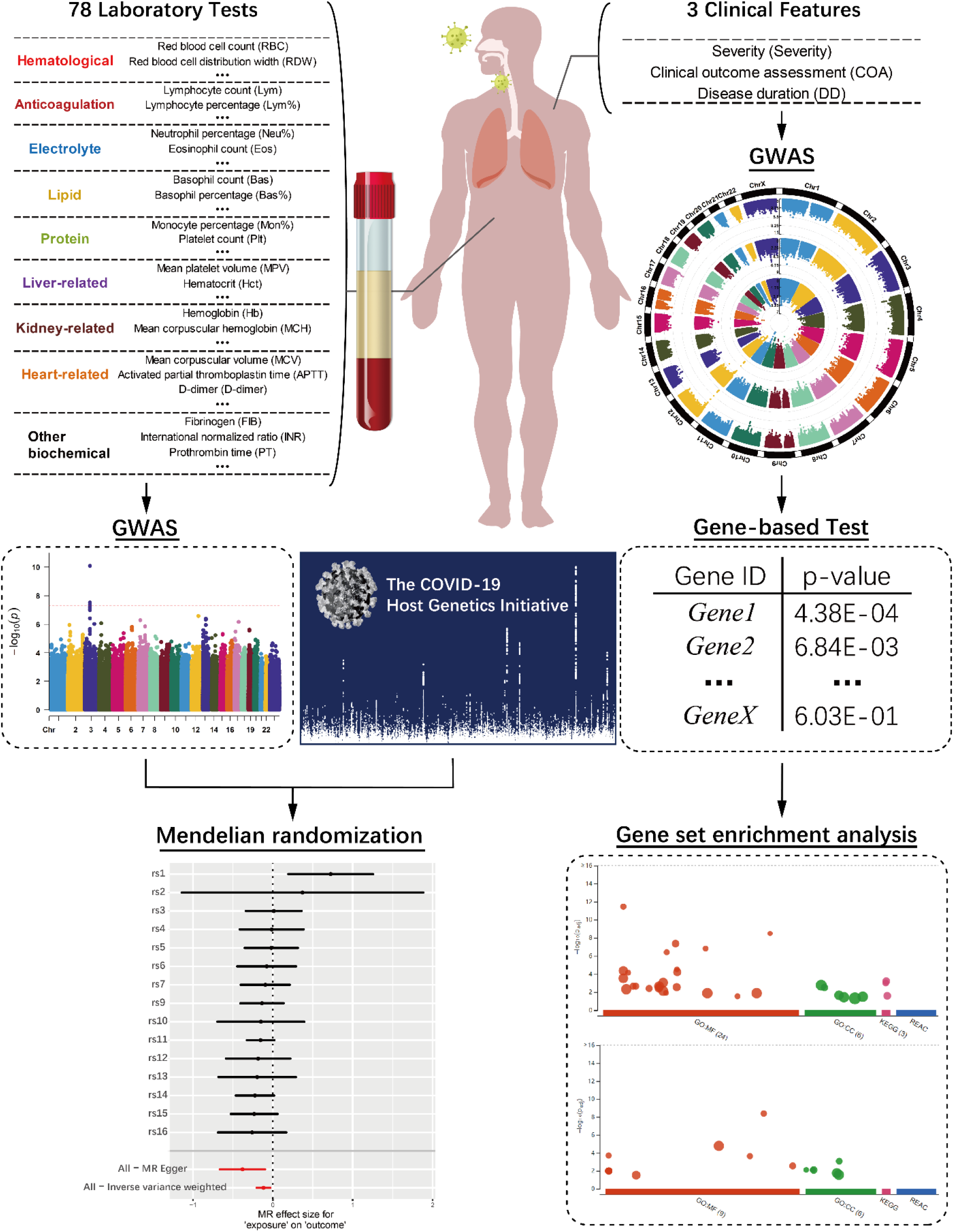
The workflow of the main analyses performed in this study

**Figure 3.**
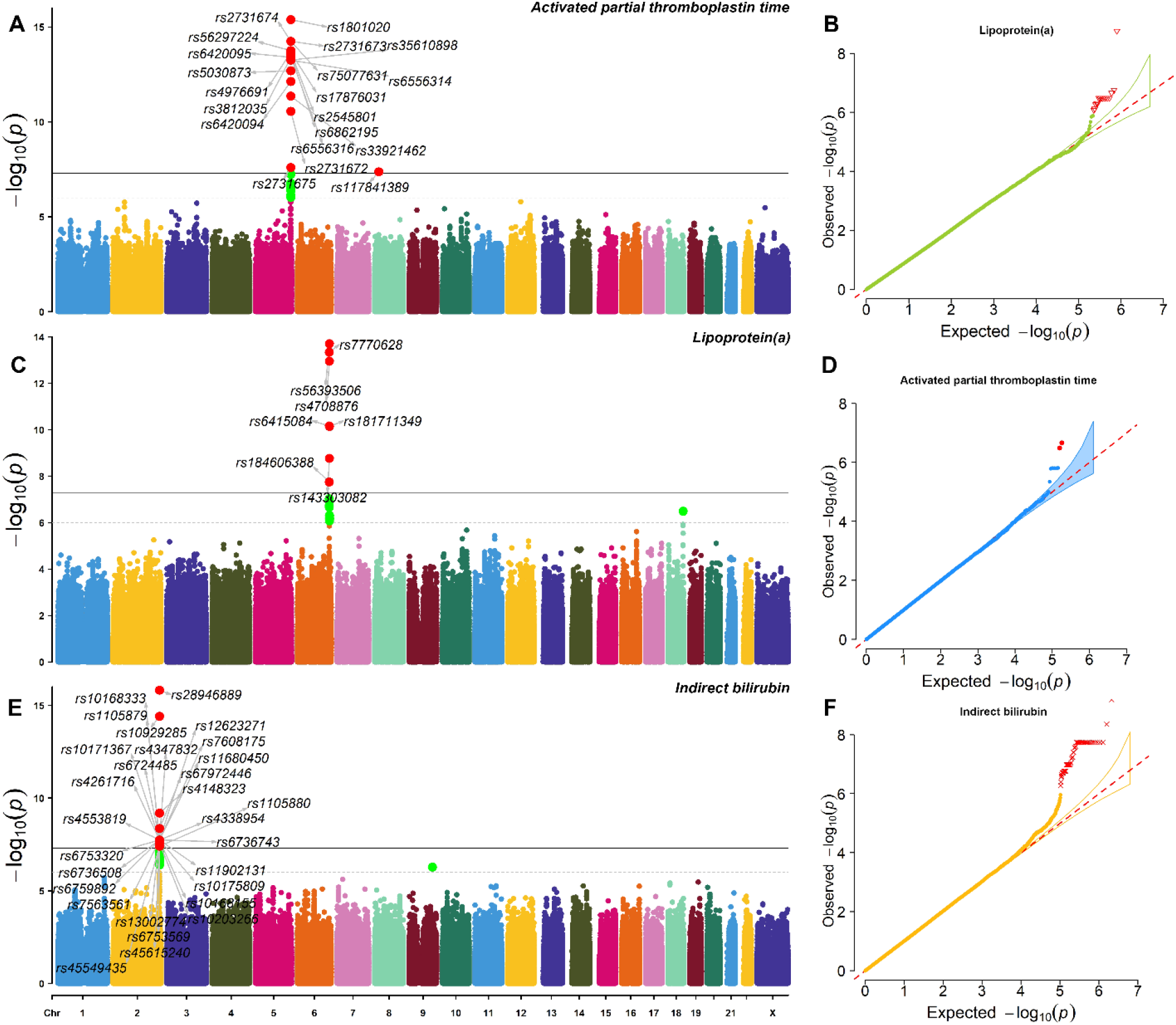
The Manhattan plots and QQ-plots of three strong signals *Notes*. Figures A, C, and E are Manhattan plots and Figures B, D, F are QQ-plots for APTT, LpA, and Ibil, respectively.

**Figure 4.**
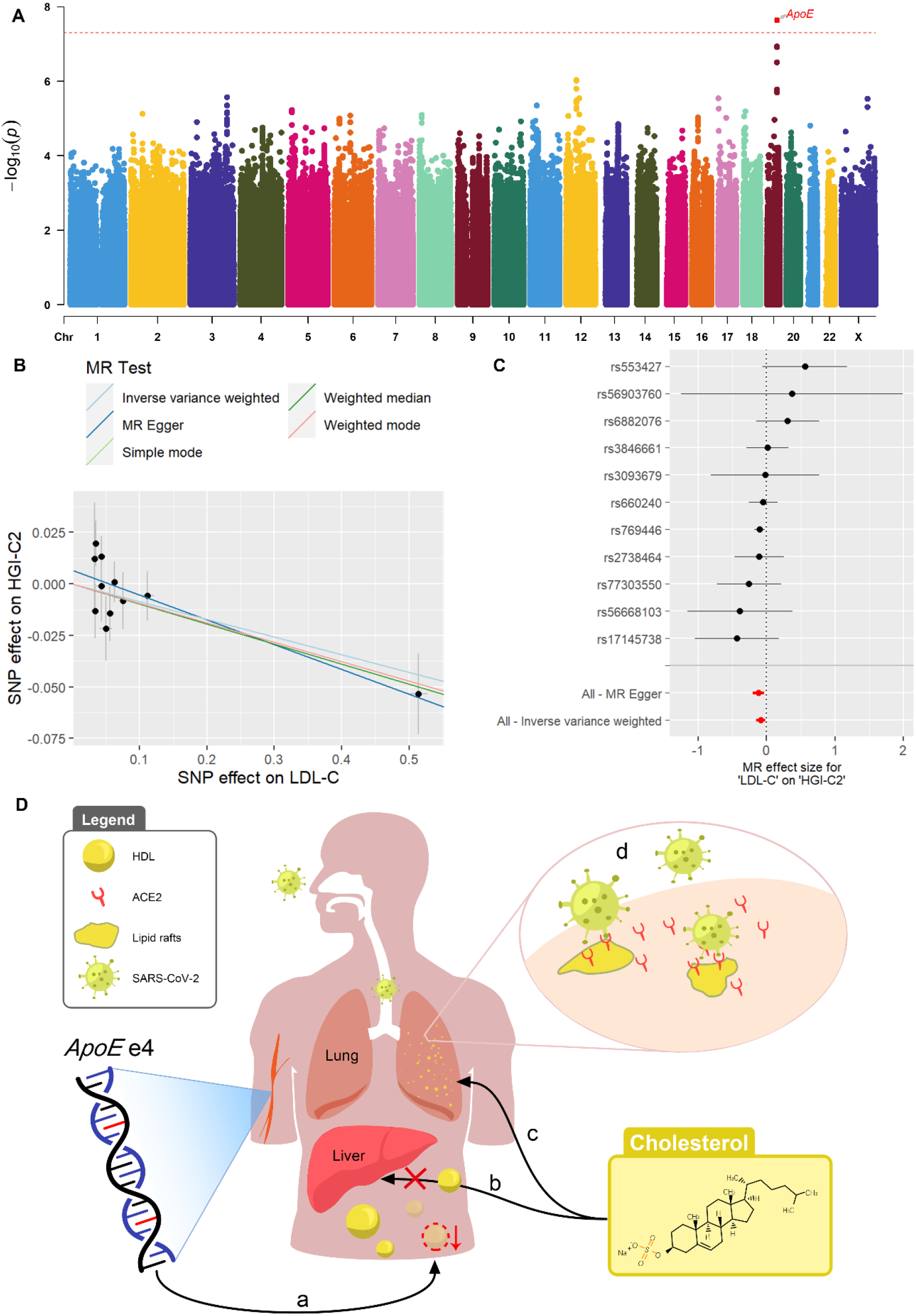
The genetic mechanisms of *ApoE* influencing severity by acting on cholesterol levels *Notes*. (A) Manhattan plot of the GWAS single-variant test results of LDL-C. The red dash line indicates the genome-wide significance threshold 5E-8. (B) The fitted line of SNP effects on severity status versus SNP effects on LDL-C with 11 SNPs significantly associated with LDL-C in BBJ database. (C) Forest plot for MR effect sizes of LDL-C on severity status based on BBJ database. (D) A genetic mechanism of how *ApoE* genotype influences the susceptibility and severity of COVID-19 disease.

**Figure 5.**
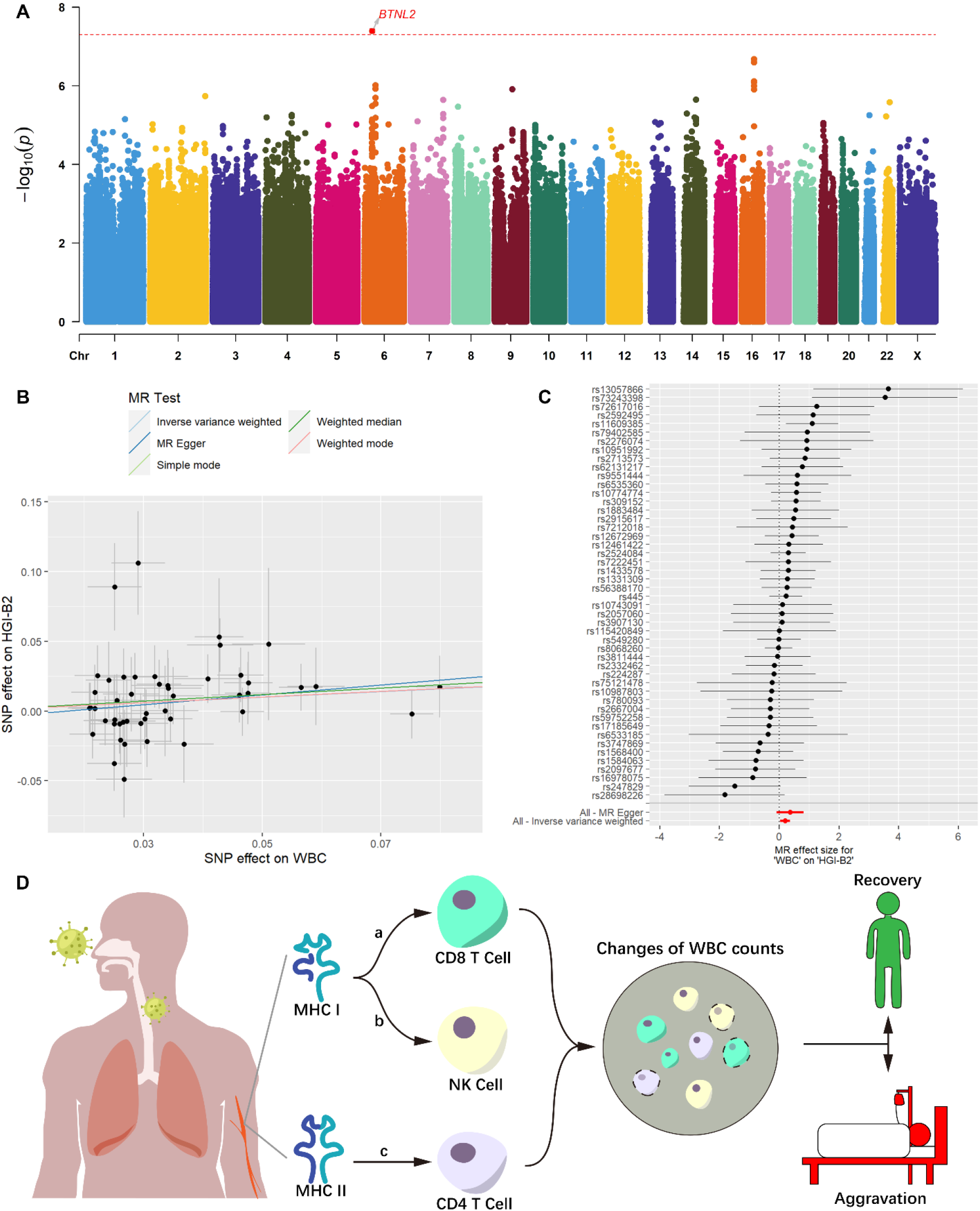
The genetic mechanisms of *MHC* system determining severity by controlling WBC counts *Notes*. (A) Manhattan plot of the GWAS single-variant test results of WBC counts. The red dash line indicates the genome-wide significance threshold 5E-8. (B) The fitted line of SNP effects on severity status versus SNP effects on WBC counts with 48 SNPs significantly associated with WBC counts in database with 151,807 East Asian participants. (C) Forest plot for MR effect sizes of WBC counts on severity status. (D) A genetic mechanism of how *MHC* system genotype influences the severity of COVID-19.

We additionally illustrated more details on the above detected associations. Specifically, the rs1801020 (*F12*)-APTT association was previously identified in GWAS analysis from the BioBank Japan Project (BBJ), one of the largest East Asian biobanks with over 160,000 subjects ^26^. The gene *F12* encodes coagulation factor XII that participates in the initiation of blood coagulation and mutation of *F12* will cause prolonged coagulation time and poor thromboplastin production ^27^. The rs56393506 (*LPA*)-LpA association was previously identified by GWAS in European population with over 13,781 individuals ^28^ but not in Asian population based on genomic studies. The gene *LPA* encodes a serine proteinase that constitutes a substantial portion of lipoprotein(a) ^29^. The rs28946889 (*UGT1A* complex)-Tbil and *UGT1A* complex-Ibil associations were identified from the BBJ database ^26^. The *UGT1A* complex represents a complex locus that encodes several UDP-glucuronosyltransferases. The mutation of *UGT1A1* gene is the only enzyme involved in bilirubin glucuronidation in hepatocytes, which can reduce the activity of the enzyme and cause insufficient bilirubin glucuronidation, thus increasing the level of serum bilirubin. The rs7412 (*ApoE*)-LDL-C association was previously identified by GWAS analysis in European, Asian, and Chinese populations. The gene *ApoE* is a type of apolipoprotein that participates in lipid metabolism and particular *ApoE* genotype results in higher risk of elevated LDL-C levels. The rs9268517-WBC is a novel genetic association identified by our GWAS analysis. However, its closest gene *BTNL2* was previously identified to be associated with WBC by a GWAS analysis with 408,112 European individuals ^30^. The gene *BTNL2* encodes MHC II type I transmembrane protein and binding to its receptor can inhibit T cell activation and cytokine production.

### Two-sample Mendelian randomization analysis

Yet we have the individual-level genotypic data, laboratory measurements, and clinical severity to perform one-sample MR analysis, we choose not to do so due to the small sample size and low powers and also its less powerful performance in controlling for confounders. We instead examined the causal relationships between the laboratory measurements with concrete genome-wide associations and COVID-19 susceptibility and severity tested by various phenotype types from the Host Genetics Initiative (HGI) database based on the two-sample MR analysis. Typically, the two-sample MR study requires two independent studies from one population to ensure consistent SNP sites. We harmonized the summary results from exposure and outcome studies to adjust the SNP sites into positive chains and remove inconsistent ones with large allele frequency difference (Supplementary Table S1). In brief, we identified three leniently significant causal associations (p-value < 0.05), including WBC counts (p-value = 0.009), LDL-C (p-value = 0.034), and apoA (p-value = 0.047) on the susceptibility and severity of COVID-19 disease. The valid instrumental variants used for LDL-C, apoA, and WBC are rs7412 (*ApoE*), rs11032789 (*EHF*), and rs9268517 (*BTNL2*), respectively. After SNPs clumping and pruning, there was only one SNP used in MR analysis for each trait and therefore the Wald ratio method ^31^ was used to estimate the causal effects. Typically, the minimum number of independent SNPs is three ^26^ and the inverse variance weighted method ^32^ is often used to estimate the causal effects. Furthermore, by controlling the false discovery rate (FDR), the q-values are all greater than 0.05 for the above identified associations with 84 multiple testing (7 laboratory traits * 12 HGI phenotypes). Despite the small number of valid instrumental variants applied in the analysis and insignificant results with large multiple testing burden, our results still provide worthwhile directions for further investigation. We detected these valuable genetic mechanisms of COVID-19 disease: *ApoE* and *MHC* family influence the COVID-19 susceptibility and severity status by acting on the cholesterol levels and WBC counts of patients, respectively.

We adopted other large-scale publicly available databases and evaluated whether our findings were also suggested by these databases. To investigate the causal effects of LDL-C, we downloaded the significant summary statistics for LDL-C from the BBJ database with sample size 72,866 ^26^. There are 22 genome-wide associated variants mapped to different genes. First, we tested on only the *ApoE* gene by using SNPs mapped to this gene (rs769446, p-value = 2.977E-322, LD with rs7412 = 0.56 in 1000 Genome Project all populations). Hereby, there was one valid instrumental SNP, and the Wald ratio test was used in MR analysis. The results were provided in Supplementary Table S2 and showed four significant associations corresponding to four HGI phenotypes. The q-values (FDR) of these associations are also less than 0.1 with two less than 0.05. Second, we did the MR analysis based on all the LDL-C associated 22 SNPs. After SNP clumping and harmonization with HGI database, 11 SNPs were used in MR. The estimate of the causal effect sizes, p-values, and q-values for testing the effect of LDL-C on susceptibility of COVID-19 were provided in Supplementary Table S3 and Figures 4B-4C. We also used MR-Egger method ^33^ to test the heterogeneity and the p-value is 0.464 > 0.05 (Q-statistic = 8.71) showing no heterogeneity. Besides the heterogeneity, we also tested pleiotropic effects based on MR-PRESSO global test ^34^ and obtained a p-value of 0.228 > 0.05 meaning no direct effects of the analyzed SNPs on outcome severity. The results of no heterogeneity and no pleiotropy enhanced the validity of MR results. We also tested the causality of cholesterol levels on COVID-19 susceptibility and severity in European population ^35^ and African American population ^36^, respectively. The results were provided in Supplementary Tables S4 and S5 showing a possible causal effect of LDL-C on COVID-19 disease in other populations besides East Asian. We mention that the causal effect directions of LDL-C on disease illness varied upon analyzed SNP(s) and HGI phenotypes; showing the complex biological mechanisms of how the genetic variation regulates the disease status by controlling cholesterol levels.

Our analysis results and observations of trait measurements over time are consistent with two main points of view. The first one is that people with *ApoE* ε4/ε4 genotype tend to have higher plasma cholesterol levels compared to those with the other *ApoE* genotypes ^37,38^. Recently, several studies reported that patients who carried *ApoE* ε4/ε4 genotype tend to be infected by SARS-CoV-2 and experience severe symptoms from COVID-19 ^39,40^. For example, a study concluded that, among older people, patients with *ApoE* ε4/ε4 genotype had much higher risk of developing severe symptoms compared with *ApoE* ε3/ε3 (OR = 2.31, p-value = 1.19E-06) ^39^. By investigating the *ApoE* genotypes in all 466 COVID-19 patients, we found 7 patients who carried *ApoE* ε4/ε4 in total, of which 5 patients were severe. We investigated the biological mechanisms of how *ApoE* gene regulated the COVID-19 susceptibility and severity and found a pathway where *ApoE* influenced disease status by controlling for cholesterol levels, which were consistent with our MR findings. Specifically, people with *ApoE* ε4/ε4 have increased risk of high cholesterol levels. When they are exposed to SARS-CoV-2, the accumulation of cholesterol in alveolar epithelial cells increased the density of lipid rafts, from which the virus binds to its target receptor ACE2. Therefore, higher density of lipid rafts facilitates the bindings in cell membranes and eventually raised the susceptibility to SARS-CoV-2 infection and severity of COVID-19^14,40,41^. The genetic mechanism is illustrated in Figure 4D. The second point of view is that as the disease condition worsened, the lipid levels including apoA and LDL-C largely decreased ^42^. Our results in the next section of “Time-series analysis of laboratory features” supported this association, showing a very low level of cholesterol in blood is a risk sign for suffering severe symptoms in COVID-19 cases.

We further tested on the *MHC* system genes. The SNPs (rs114398276) mapped to *MHC* genes in BBJ database were removed when harmonizing with HGI results due to large difference in allele frequency. As an alternative, we downloaded another summary result based on East Asian population with sample size 151,807 ^43^. First, we tested the causality of WBC counts on COVID-19 severity based on SNPs mapped to *MHC* family, which is gene *HLA-C* (rs2524084, p-value = 1.260E-53, LD with rs9268517 = 0.21 in 1000 Genome Project all populations). The results were provided in Supplementary Table S6 and showed four significant associations corresponding to four HGI phenotypes. The q-values (FDR) of these four associations are also less than 0.05 suggesting the candidate pathway that *MHC* family has an effect on COVID-19 disease by controlling WBC counts. Second, we did the MR analysis based on all associated 81 SNPs. After SNPs clumping and harmonization with HGI database, 48 SNPs were used in MR. The estimates of the effect size, p-values, and q-values for testing the causal effects of WBC counts on disease illness were provided in Supplementary Table S7 and Figures 5B-5C. We also used MR-Egger method to test the heterogeneity and the p-value is 0.365 > 0.05 (Q-statistic = 48.69) showing no heterogeneity. Besides the heterogeneity, we also tested pleiotropic effects and obtained a p-value of 0.469 > 0.05 meaning no pleiotropy. The causal effect direction of WBC counts on COVID-19 illness is consistently positive based on our dataset and the tested Asian database, suggesting that WBC count is likely a risk predictor to disease status.

We searched the gene-trait “major histocompatibility complex” + COVID-19 on PubMed and it yielded 57 hits (as of May 10, 2021), indicating the essential role of the *MHC* system in the immune responses of COVID-19 patients. The MHC complex is a group of related proteins that are encoded by the *MHC* gene complex in human. The function of these cell-surface proteins is to activate T lymphocyte cells and NK (natural killer) cells by presenting antigens. The SARS-CoV-2 was found to restrain antigen presentation and suppress immune reaction by downregulating the expression of *MHC* class in COVID-19 cases ^44^. Previous studies showed that, as the disease progresses, mHLA-DR levels and lymphocyte cell counts varied in COVID-19 patients ^15,45^. In conclusion, the *MHC* gene complex and its expression levels are closely related to COVID-19 severity of infection symptoms. The genetic mechanism is illustrated in Figure 5D.

### Reverse Mendelian randomization analysis

Our MR analysis in previous section suggested the causal effects of LDL-C and WBC counts on COVID-19 disease. To further investigate whether there exist causal effects of COVID-19 severity on LDL-C or WBC counts, we did the reverse MR analysis where the illness status was exposure, and the laboratory assessments were outcome variables. Specifically, we used the various HGI phenotypes ^9^ as exposure and LDL-C levels and WBC counts as outcome. The results were provided in Supplementary Tables S8 and S9 implying no causality.

### Time-series analysis of laboratory features

We used line chart to visualize the variation patterns of laboratory measurements at five time points. Based on the severity status at admission and clinical outcome in the end, the patients were grouped into three teams: mild and recovered (mild), severe and recovered (recovery), and severe and dead (death). We provided the line charts for apoA, LDL-C, and WBC in Figures 6A-6C and for the other traits in Supplementary Figures. These line charts could visually reflect whether a trait was protective or risk to COVID-19 disease. For example, the apoA and LDL-C levels were likely to be protective factors since patients in the recovery and death groups had lower values than those in mild group, while the WBC counts appeared to be risk predictor to COVID-19. We also performed logistic regression analysis to statistically assessed the risk and protective factors to disease severity and clinical outcome (Figures 6D-6F). For apoA and LDL-C, negative effects implied protective influence on disease illness; while for WBC counts, the effect direction is positive indicating risk impact. We also noticed that with the progress of the disease, the negative association between the lipid traits and clinical outcome became more significant. Considering that patients in mild and recovery groups were cured and out of hospital, these time-series analyses also could reflect the comparison of laboratory features between COVID-19 cases and healthy controls.

**Figure 6.**
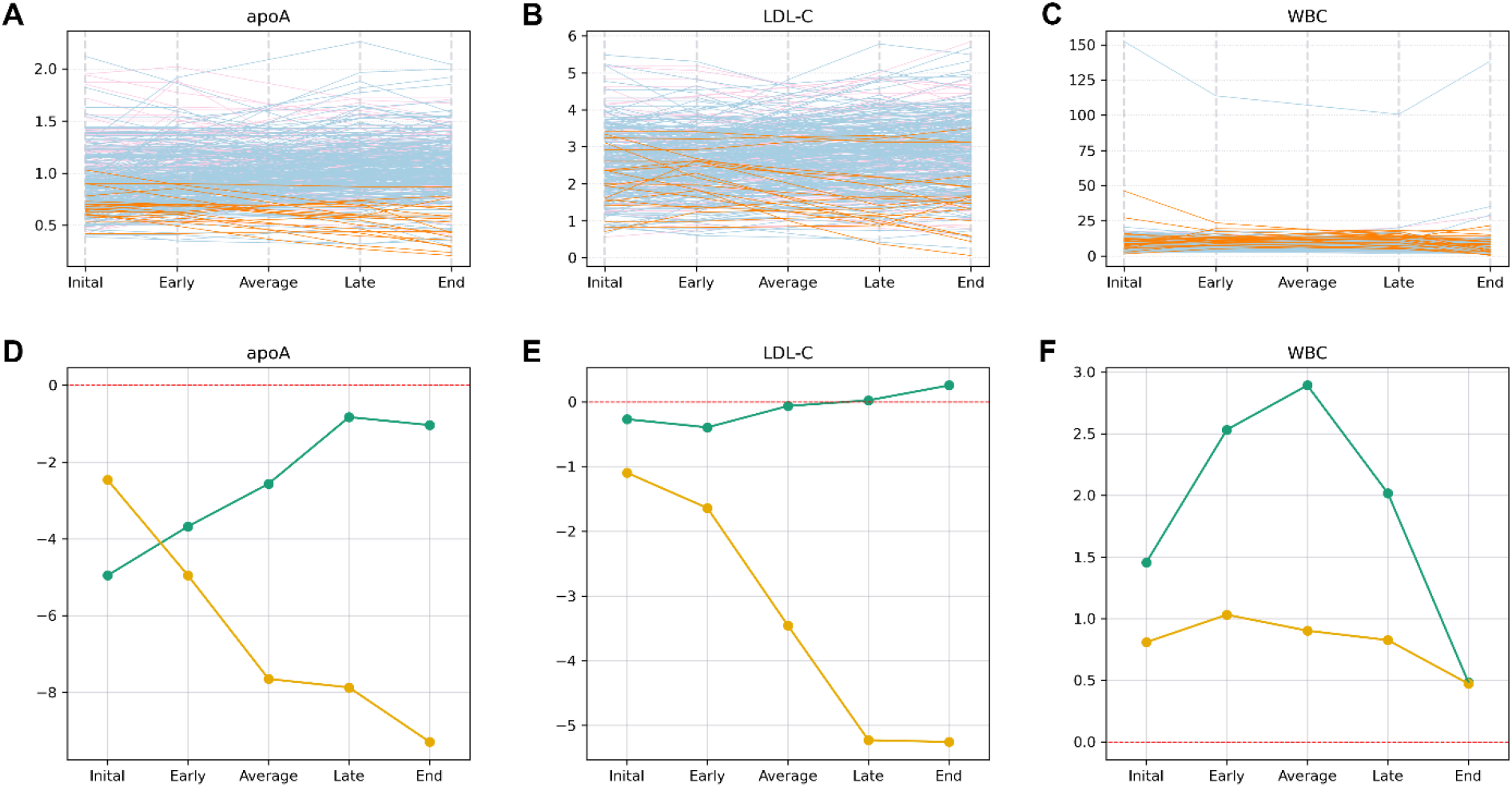
The time-series laboratory features *Notes*. The top three figures show variation trends of apoA, LDL-C, and WBC counts over time. The pink, light blue, and orange lines indicate the patients are grouped into mild, recovery, and death team, respectively. The y-axis denotes the quantities of each trait. The bottom three figures show regression associations between each trait with disease severity (green) and clinical outcome (yellow)over time. The y-axis denotes -log10(p-value) multiplied by the effect direction (positive effect is 1 and negative effect is -1).

### Gene-based and gene-set enrichment analysis of clinical measurements

We analyzed three clinical features, including severity (mild versus severe), clinical outcome assessments (survival versus death), and disease duration (hospitalized days) by first performing single-variant genome-wide association studies. The Circular-Manhattan plot and QQ-plot were provided in Figures 7A-7B. No genetic variants reach the genome-wide significance threshold (p-value < 5E-08) due to the current small sample size (*N* = 466) and thus the effect sizes of single variants tend to be small. To aggregate the single-variant effects, we further performed VEGAS gene-based test ^46^ and g:GOST GSEA analysis ^47^ for clinical severity. With a window size of 50kb, 25,345 genes were mapped and the average number of SNPs on each gene is 251. For the window of 10kb, 24,640 genes were mapped, and the average number of SNPs is 119. Then, we selected only genes with p-value less than 0.05. A number of 1,170 genes passed the significance threshold for window size 50kb and 1099 genes for 10kb. We obtained an intersect of 705 genes from the two sets of significant genes for further GSEA analysis.

**Figure 7.**
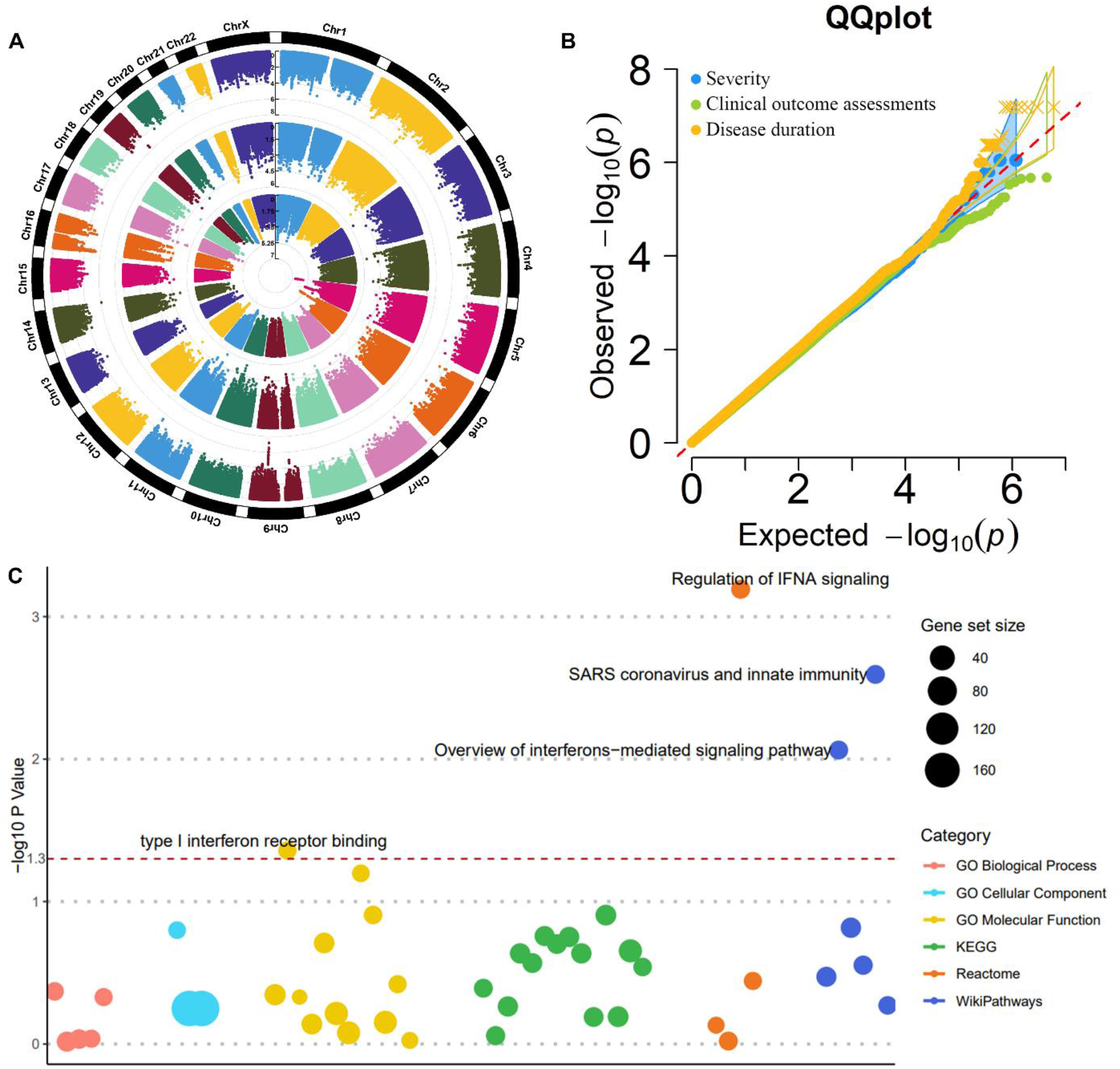
The genome-wide association studies of COVID-19 severity: a case-control study *Notes*. (A) Circular-Manhattan plots for clinical traits. The inner circle is for severity status (mild vs. severe), the middle circle is for clinical outcome assessments (survival vs. death), and the outer circle is for disease duration (hospitalized days). (B) The QQ-plot for three clinical traits. (C) Bubble plot of GSEA analysis based on the single-variant and gene-based studies on severity status. The red dashed line is the threshold of 0.05.

The GSEA results identified four significant pathways with p-value less than 0.05 (Figure 7C). These pathways include regulation of IFNA signaling (REAC:R-HSA-912694, p-value = 6.42E-04), SARS coronavirus and innate immunity (WP:WP4912, p-value = 2.54E-03), overview of interferons-mediated signaling pathways (WP:WP4558, p-value = 8.64E-03), and type I interferon receptor binding (GO:0005132, p-value = 4.38E-02). All the four pathways belong to the IFNA family, which is a member of the alpha interferon gene cluster on and encodes type I interferon (IFN) family produced in response to viral infection. The IFNA family is a key part of the innate immune response with potent antiviral, antiproliferative and immunomodulatory properties. Insufficient virus-induced type I IFN production is characteristic of SARS-CoV-2 infection since SARS-CoV-2 suppresses the IFN response by interacting with essential IFN signaling pathways ^48^. Blunted amounts of IFNs have been detected in the peripheral blood or lungs of severe COVID-19 patients ^49^. We note that since VEGAS is based on a simulation procedure to calculate the gene-based p-values, thus its results may vary slightly every time we rerun the analysis. We examined the effect of running the analysis multiple times and found that the results of gene-based association and the subsequent GSEA study are robust and reliable. We also investigated the effect of varying the window sizes around each gene and found that the results are robust to the choice of window sizes. In summary, based on the single variant associations, VEGAS gene-based tests, and GSEA analysis, we identified four IFNs pathways whose imbalanced responses may cause the pathology of COVID-19 based on genomic studies in Chinese population.

As we mentioned in the Introduction section, several genetic loci have been identified to be associated with the critical illness in COVID-19 ^7^. We summarized eight genome-wide significant associations in Table 3, including the lead SNP in each locus, the p-values of these SNPs for testing severity status in our dataset, and their allele frequencies in Asian and European populations from 1000 Genome Project and in our case subjects. Among these eight SNPs, one SNP (rs74956615, 19:10427721) does not exist in our imputed genotype and two SNPs (rs73064425, 3:45901089; rs3131294, 6:32180146) were removed from analysis due to low allele frequencies. For four out of the other five SNPs, their allele frequencies in European populations are much higher than in Asian population (average difference is 0.16), showing that these significant SNPs are more prominent in European than in Asian. The eighth SNP is rs9380142 (6:29798794) mapped to gene *HLA-G*. The *HLA-G* gene belongs to the MHC region that plays a critical role in immune responses and regulations. We believe that a large-scale COVID-19 case-control study in Chinese population also has potential to uncover the MHC region.

**Table 3.** The summary of eight reported COVID-19 illness associated loci

| SNP | Chr:BP | Locus | p-value | A1 | 1KGP Asian Freq | 1KGP European Freq | A1 Freq | Mild Freq | Severe Freq |
| --- | --- | --- | --- | --- | --- | --- | --- | --- | --- |
| rs73064425 | 3:45901089 | LZTFL1 | / | T | 0.005 | 0.0795 | 0.005365 | 0.005882 | 0.005068 |
| rs9380142 | 6:29798794 | HLA-G | 0.846804 | G | 0.3492 | 0.3439 | 0.43133 | 0.4206 | 0.4375 |
| rs143334143 | 6:31121426 | CCHCR1 | 0.966751 | A | 0.0347 | 0.1123 | 0.059013 | 0.06176 | 0.05743 |
| rs3131294 | 6:32180146 | NOTCH4 | / | A | 0.0079 | 0.1133 | 0.006438 | 0.008824 | 0.005068 |
| rs10735079 | 12:113380008 | OAS1–<br>OAS3 | 0.433044 | G | 0.252 | 0.3638 | 0.233906 | 0.2265 | 0.2382 |
| rs2109069 | 19:4719443 | DPP9 | 0.788196 | A | 0.1399 | 0.3211 | 0.143777 | 0.1382 | 0.147 |
| rs74956615 | 19:10427721 | TYK2 | / | / | / | / | / | / | / |
| rs2236757 | 21:34624917 | IFNAR2 | 0.4025 | G | 0.4345 | 0.7058 | 0.381974 | 0.3618 | 0.3936 |

*Notes.* A1 denotes the effect/alternate allele. 1KGP Asian Freq indicates the allele frequency in Asian population from the 1000 Genome Project. Mild Freq indicates the allele frequency in patients from mild and moderate (grouped into mild) groups.

## Materials and methods

### Subjects

All the subjects enrolled in this study were recruited by the Wuhan Union Hospital (Union hospital of Tongji Medical College of Huazhong University of Science and Technology). These subjects had been diagnosed with COVID-19 respiratory disease and hospitalized in Wuhan Union Hospital between January 15 and April 4, 2020. Written informed consent was obtained from all participants, as approved by the Medical Ethics Committee of Union Hospital, Tongji Medical College, Huazhong University of Science and Technology.

### Phenotype

There are two types of phenotypes: laboratory and clinical measurements. Numerous laboratory features from various lab test categories were measured at different time points during hospitalization. For each laboratory measurement, we took average of all non-missing records of each patient during his or her hospitalization for genomic analysis. The clinical characteristics include three traits: severity status (mild versus severe) collected at the time of admission to the hospital, clinical outcome assessments (survival versus death), and disease duration (i.e., hospitalized days) at the time of eventual treatment and prevention of disease.

### Genotyping and imputation

We sequenced samples with the DNBSEQ platform (MGI, Shenzhen, China) to generate 100bp paired-end reads. The mean sequencing depth was 17.8×. We excluded samples with (i) sample call rate < 0.99, (ii) closely related individuals identified by identity-by-descent (IBD > 0.1) calculated in KING ^50^, and (iii) outliers identified by principal component analysis based on three-sigma rules. We then applied standard quality control criteria for genetic variants by removing those with (i) SNP call rate < 0.99, (ii) minor allele frequency (MAF) < 0.01, and (iii) Hardy-Weinberg equilibrium p-value < 1E-06. Based on the VCF files after VQSR with biallelic variants, imputation was performed with Beagle v4.0 ^51^ taking GL as input in east Asian (EAS) population of 1000 Genomes Project as reference panel.

### Genome-wide association studies

We used PLINK v2.0 ^52^ to perform single-variant GWAS analyses using a linear regression model for the quantitative laboratory features under the assumption of additive allelic effects of the SNP dosage. For each trait, we adjusted for age, sex, and top six principal components (PCs) of genetic ancestry and normalized the resulting residuals by applying a Z-score normal transformation. The number of PCs was chosen by using EIGENSTRAT software ^53,54^. We set a genome-wide significance threshold at the level of 5E-08 and a study-wide significance threshold at the level of 6.41E-10 (=5E-08/78) by applying Bonferroni correction based on the number of laboratory traits (n = 78).

### Two-sample Mendelian randomization

Several significant associations were identified from the GWAS analysis of laboratory measurements. Given the potential genetic correlation between these features and the COVID-19 susceptibility and severity, we performed two-sample Mendelian randomization analyses to examine causal effects between them and uncover genetic variants that determined disease status by acting on the laboratory traits. Note that the causal interpretation of the laboratory exposure variable on the disease outcome requires three standard assumptions to hold: (i) relevance: instrumental variants are highly associated with the exposure; (ii) no unmeasured confounders: variants are not associated with any confounding factors that may be associated with both exposure and outcome; and (iii) exclusion restriction: variants influence the outcome only through the path of exposure, i.e., no horizontal pleiotropic effects of variants on the outcome. We used the laboratory features that displayed strong study-wide signals with genetic variants (p-value < 6.41E-10) as the variant-exposure associations to ensure the relevance assumption. For the variant-outcome associations, we used the COVID-19 Host Genetics Initiative (HGI) round 5 GWAS meta-analysis results and genome-wide significant variants were removed to ensure the exclusion restriction assumption. There are four types of phenotypes in HGI: very severe respiratory confirmed covid versus population (A2), hospitalized covid versus not hospitalized covid (B1), hospitalized covid versus population (B2), and covid versus population (C2). We selected B2, C2, and B1 phenotypes to study the susceptibility and critical illness of COVID-19. For each type of phenotype, there are four different sets of populations: all populations but not 23andme, all populations but not UKBB, all Europeans, and all Europeans but not UKBB. We used R (version 4.0.2) with the *TwoSampleMR* package ^55,56^ and set the significance threshold at the level of 0.05. We also calculated the *adjusted* p-values (i.e., q-values) by controlling the false discovery rate (FDR). In details, we used R (version 4.0.2) with the p.adjust function from *stats* package ^57^ to obtain the q-values and declared more stringently significant associations based on an FDR of 0.1 and 0.05.

### Time-series laboratory features

To better understand the variation trends of laboratory features during the patients’ hospital stay, we divided the hospitalization days into several stages for each patient. For each laboratory feature, we only kept patients with more than two non-missing records and divided these records into two equal-length groups named early and late time stage. At each of the two-time stages, we took average of all available values for each feature and treated this average as the patient’s representative measurement at this time stage. We also defined the first and last non-missing measurement as initial and end record. In addition, we took average of all non-missing records for each patient during his or her hospitalization days and used this value as the overall average. By doing so, for each patient, we obtained five values of all laboratory features: initial, early, average, late, and end. To test the association between the time-series laboratory traits with disease severity and clinical outcome, we further performed logistic regression analysis after adjusting for age and sex at each time stage.

### Gene-based and gene-set enrichment analysis

For the patients’ clinical features, we performed GWAS single-variant analysis in PLINK 2.0 based on a logistic (for severity status and clinical outcome assessments) or linear (disease duration) regression model. For the severity status, we further conducted gene-based tests and gene-set enrichment analysis (GSEA) to aggregate effects of multiple genetic variants from the single tests. The gene-based test is VEGAS method ^46^ that combines the p-values of the single variants. A list of selected genes from the gene-based results was taken for further GSEA analysis to uncover functional pathways based on the g:GOST toolset ^47^. We used six existing gene set databases, including GO (gene ontology) molecular function ^58^, GO cellular component ^58^, GO biological process ^58^, KEGG (Kyoto encyclopedia of genes and genomes) ^59^, Reactome ^60^, and WikiPathways ^61^.

### Data availability

The data that support the findings of this study have been deposited into CNGB Sequence Archive (CNSA) ^62^ of China National GeneBank DataBase (CNGBdb) ^63^ with accession number CNP0001876.

## Discussion

The severe acute respiratory syndrome coronavirus 2 (SARS-CoV-2) is a new coronavirus causing the ongoing pandemic coronavirus disease 2019 (COVID-19). Patients’ with COVID-19 experience largely various clinical and laboratory assessments, from no symptoms, to exhausted respiratory system, and even death. Many clinical and experimental studies have concluded that several key determinants are responsible for the disease variability, including old age, male gender, and having comorbidities at the admission to the hospital. However, these factors still cannot fully account for the diverse symptoms among patients. Recent studies have turned more attentions into the host genetic background. The Host Genetics Initiative (HGI) have reported many candidate loci by performing large-scale GWAS analysis with thousands of cases and up to millions of controls.

In this study, we analyzed 466 COVID-19 patients hospitalized in the Wuhan Union Hospital. A broad range of clinical information, such as age, gender, comorbidities, and laboratory blood test results, such as hematological and liver-related assessments were collected for each patient. The analyses of age, gender, and comorbidities in mild and severe patients confirmed their potential risk on critical symptoms. We also performed GWAS analysis for the numerous laboratory features and discovered seven concrete genome-wide variant-trait associations, five of which were previously uncovered by large-scale genomic studies. Our results were either the first replication or the first identification study in Chinese population based on GWAS study. With these well-established genetic associations, we conducted Mendelian randomization (MR) analysis to uncover important laboratory traits that have causal effects on the susceptibility and severity of COVID-19 disease. Our analyses highlighted two fundamental pathways; one is the cholesterol levels with functional gene *ApoE*, and the other one is the white blood cell counts (WBC) with functional gene *MHC* complex. We further researched on and well explained the genetic mechanisms of how genes *ApoE* and *MHC* family influenced the disease status by acting on cholesterol levels and WBC counts.

We additionally carried out the gene-based tests and gene-set enrichment analysis (GSEA) based on the single-variant GWAS summary results of severity case-control study. Interestingly, we for the first time revealed four interferons (IFNs) related functional pathways based on host genetic studies in Chinese population, including regulation of IFNA signaling, SARS coronavirus and innate immunity, overview of interferons-mediated signaling pathway, and type I interferon receptor binding. Several considerable studies observed that low levels of IFNs production was highly correlated with severe COVID-19. Most of these studies were based on bulk RNA-seq, scRNA-seq, or experimental designs, while our analysis is built on genomic data, supporting this solid conclusion from a new perspective.

Despite the many compelling discoveries of our work, there are still a few limitations. First, the single-variant GWAS analysis of severity status did not identify any genome-wide signals due to the current sample size (*N* = 466) and thus small genetic effect sizes. We believe that large-scale case-control studies have potentials to uncover genome-wide significant variants. Second, even though our GWAS analysis of laboratory features produced concrete and powerful signals, there are still many more traits without being detected associated variants that reach the study-wide or the genome-wide significance threshold. For the identified associations, after SNPs clumping and pruning, there is merely one independent strong variant, while the tested traits were often known as polygenic. This is still due to small sample size restriction. Third, despite the fact that our MR findings are supported by solid biological mechanisms and also potentially replicated by many populations, the causal significance could be different among different HGI phenotypes. For example, when testing the causal effects of LDL-C based on BBJ database, with the outcome of covid vs. population in all population without UKBB, the p-value is 0.01; while for the outcome of hospitalized covid vs. population in all population without UKBB, the p-value is 0.98. We consider this phenomenon directly relating to the corresponding HGI GWAS summary results and further investigations are needed to explain the intrinsic biological reasons.

## Data Availability

The data that support the findings of this study have been deposited into CNGB Sequence Archive (CNSA) of China National GeneBank DataBase (CNGBdb) with accession number CNP0001876.

## Author contribution

F.C. and X.J. conceived the study, designed the research program and managed the project.

F.Z., Y.J., Z.L., L.T., N.X., D.H., X.Z., R.X., Y.C., and W.L. collected the samples.

Y.L. finished the laboratory processing and data acquisition.

Z.L. and P.L. preprocessed the data and finished the quality control.

H.Z., L.L. and J.Z. performed the statistical analyses.

Y.S., S.L., and X.S. advised on statistical methods.

H.Z, F.Z, L.L., Y.J., Y.L. Z.L. and J.Z. wrote the manuscript.

All authors participated in revising the manuscript.

## Acknowledgement

This study was supported by National Natural Science Foundation of China (32000398), Natural Science Foundation of Guangdong Province, China (2017A030306026), Guangdong-Hong Kong Joint Laboratory on Immunological and Genetic Kidney Diseases (2019B121205005), The Innovative Major Emergency Project Funding against the COVID-19 in Hubei Province (No. 2020FCA041), the Innovative Major Emergency Project Funding against the COVID-19, HUST (No. 2020kfyXGYJ039), Guangdong Provincial Key Laboratory of Genome Read and Write(No. 2017B030301011) and China National GeneBank.

