## Supplementary material for "A Chinese host genetic study discovered type I interferons and causality of cholesterol levels and WBC counts on COVID-19 severity": Supplementary tables-Xiehe.COVID-19.pdf

S1.Xiehe lab as exposure

| outcome | exposure | method | nsnp | b | se | p-value | q-value |
|---|---|---|---|---|---|---|---|
| B2_ALL_eu | WBC_coun | Wald ratio | 1 | 0.156613 | 0.059917 | 0.008953 | 0.752068 |
| C2_ALL_eu | WBC_coun | Wald ratio | 1 | 0.079034 | 0.035063 | 0.024192 | 0.762951 |
| B1_ALL_lea | LDL | Wald ratio | 1 | 0.170973 | 0.080656 | 0.034023 | 0.762951 |
| B1_ALL_eu | apoA | Wald ratio | 1 | 0.108992 | 0.054961 | 0.047357 | 0.762951 |
| B2_ALL_lea | WBC_coun | Wald ratio | 1 | 0.091528 | 0.047762 | 0.055321 | 0.762951 |
| B2_ALL_lea | LDL | Wald ratio | 1 | 0.085008 | 0.045677 | 0.062734 | 0.762951 |
| C2_ALL_lea | WBC_coun | Wald ratio | 1 | 0.051371 | 0.027692 | 0.063579 | 0.762951 |
| C2_ALL_eu | apoA | Wald ratio | 1 | 0.018017 | 0.010557 | 0.087909 | 0.923044 |
| B1_ALL_lea | LDL_A | Wald ratio | 1 | 0.115748 | 0.071338 | 0.104691 | 0.977111 |
| B1_ALL_eu | LDL | Wald ratio | 1 | 0.13349 | 0.090469 | 0.140072 | 0.99301 |
| C2_ALL_eu | apoA | Wald ratio | 1 | 0.018368 | 0.012738 | 0.14931 | 0.99301 |
| C2_ALL_lea | LDL_A | Wald ratio | 1 | -0.02009 | 0.015063 | 0.182298 | 0.99301 |
| B1_ALL_lea | LDL_A | Wald ratio | 1 | 0.065019 | 0.049606 | 0.189954 | 0.99301 |
| B1_ALL_eu | LDL_A | Wald ratio | 1 | 0.111567 | 0.091336 | 0.221898 | 0.99301 |
| B1_ALL_lea | LDL | Wald ratio | 1 | 0.126633 | 0.107545 | 0.239002 | 0.99301 |
| B2_ALL_eu | WBC_coun | Wald ratio | 1 | 0.079103 | 0.06721 | 0.239213 | 0.99301 |
| B1_ALL_eu | apoA | Wald ratio | 1 | 0.041002 | 0.034992 | 0.241294 | 0.99301 |
| C2_ALL_lea | LDL_A | Wald ratio | 1 | -0.02068 | 0.017757 | 0.244293 | 0.99301 |
| B1_ALL_eu | WBC_coun | Wald ratio | 1 | 0.117882 | 0.105119 | 0.262114 | 0.99301 |
| B2_ALL_eu | LDL_A | Wald ratio | 1 | 0.046309 | 0.044332 | 0.2962 | 0.99301 |
| C2_ALL_eu | LDL | Wald ratio | 1 | 0.028225 | 0.027071 | 0.29712 | 0.99301 |
| B2_ALL_eu | LDL | Wald ratio | 1 | 0.051399 | 0.05281 | 0.330407 | 0.99301 |
| B2_ALL_eu | APTT | Wald ratio | 1 | -0.04491 | 0.047806 | 0.347561 | 0.99301 |
| B2_ALL_lea | APTT | Wald ratio | 1 | -0.035 | 0.038022 | 0.357284 | 0.99301 |
| B2_ALL_lea | apoA | Wald ratio | 1 | -0.01823 | 0.019818 | 0.357769 | 0.99301 |
| B1_ALL_eu | LDL_A | Wald ratio | 1 | 0.051335 | 0.056443 | 0.363082 | 0.99301 |
| B1_ALL_eu | APTT | Wald ratio | 1 | -0.07248 | 0.085623 | 0.397278 | 0.99301 |
| B1_ALL_lea | apoA | Wald ratio | 1 | 0.039981 | 0.04799 | 0.404785 | 0.99301 |
| C2_ALL_lea | WBC_coun | Wald ratio | 1 | 0.025773 | 0.031156 | 0.408111 | 0.99301 |
| B2_ALL_eu | apoA | Wald ratio | 1 | 0.022877 | 0.027781 | 0.410251 | 0.99301 |
| C2_ALL_lea | apoA | Wald ratio | 1 | 0.007764 | 0.009721 | 0.424477 | 0.99301 |
| C2_ALL_lea | LDL | Wald ratio | 1 | 0.019264 | 0.024565 | 0.432911 | 0.99301 |
| B2_ALL_lea | LDL | Wald ratio | 1 | 0.039791 | 0.052188 | 0.445785 | 0.99301 |
| C2_ALL_eu | WBC_coun | Wald ratio | 1 | 0.031171 | 0.041034 | 0.447461 | 0.99301 |
| B1_ALL_lea | Ibil | Wald ratio | 1 | 0.064846 | 0.086171 | 0.451737 | 0.99301 |
| B1_ALL_lea | Tbil | Wald ratio | 1 | 0.072831 | 0.096783 | 0.451737 | 0.99301 |
| B2_ALL_lea | WBC_coun | Wald ratio | 1 | 0.037393 | 0.052203 | 0.473806 | 0.99301 |
| C2_ALL_eu | APTT | Wald ratio | 1 | -0.01516 | 0.022094 | 0.492714 | 0.99301 |
| C2_ALL_lea | apoA | Wald ratio | 1 | 0.007694 | 0.011494 | 0.503269 | 0.99301 |
| B2_ALL_eu | LDL_A | Wald ratio | 1 | 0.022833 | 0.035163 | 0.516116 | 0.99301 |
| B1_ALL_lea | WBC_coun | Wald ratio | 1 | 0.05349 | 0.084908 | 0.528712 | 0.99301 |
| B1_ALL_lea | APTT | Wald ratio | 1 | -0.04161 | 0.067834 | 0.539566 | 0.99301 |
| B2_ALL_eu | LDL | Wald ratio | 1 | -0.03825 | 0.062633 | 0.541358 | 0.99301 |
| C2_ALL_lea | APTT | Wald ratio | 1 | -0.00973 | 0.01627 | 0.549623 | 0.99301 |
| C2_ALL_eu | APTT | Wald ratio | 1 | -0.01059 | 0.018509 | 0.56735 | 0.99301 |
| C2_ALL_lea | APTT | Wald ratio | 1 | -0.01042 | 0.018817 | 0.579778 | 0.99301 |
| B2_ALL_lea | apoA | Wald ratio | 1 | -0.01095 | 0.023903 | 0.646995 | 0.99301 |
| B2_ALL_lea | APTT | Wald ratio | 1 | -0.01483 | 0.032552 | 0.648679 | 0.99301 |

| | | | | | | | |
|---|---|---|---|---|---|---|---|
| B2_ALL_eu | Ibil | Wald ratio | 1 | -0.01874 | 0.041973 | 0.655293 | 0.99301 |
| B2_ALL_eu | Tbil | Wald ratio | 1 | -0.02104 | 0.047141 | 0.655293 | 0.99301 |
| B1_ALL_eu | Ibil | Wald ratio | 1 | 0.038356 | 0.099885 | 0.700974 | 0.99301 |
| B1_ALL_eu | Tbil | Wald ratio | 1 | 0.04308 | 0.112185 | 0.700974 | 0.99301 |
| B2_ALL_eu | apoA | Wald ratio | 1 | 0.008296 | 0.022173 | 0.708297 | 0.99301 |
| B1_ALL_eu | Ibil | Wald ratio | 1 | -0.02369 | 0.065211 | 0.716419 | 0.99301 |
| B1_ALL_eu | Tbil | Wald ratio | 1 | -0.0266 | 0.073242 | 0.716419 | 0.99301 |
| B2_ALL_eu | Ibil | Wald ratio | 1 | -0.01842 | 0.052427 | 0.725333 | 0.99301 |
| B2_ALL_eu | Tbil | Wald ratio | 1 | -0.02069 | 0.058883 | 0.725333 | 0.99301 |
| B2_ALL_eu | APTT | Wald ratio | 1 | -0.0136 | 0.038745 | 0.725618 | 0.99301 |
| C2_ALL_eu | LDL_A | Wald ratio | 1 | -0.00588 | 0.016838 | 0.727002 | 0.99301 |
| C2_ALL_lea | Ibil | Wald ratio | 1 | -0.00685 | 0.021038 | 0.744567 | 0.99301 |
| C2_ALL_lea | Tbil | Wald ratio | 1 | -0.0077 | 0.023629 | 0.744567 | 0.99301 |
| B1_ALL_eu | WBC_coun | Wald ratio | 1 | 0.045377 | 0.147615 | 0.758539 | 0.99301 |
| B1_ALL_lea | apoA | Wald ratio | 1 | 0.009377 | 0.032476 | 0.77277 | 0.99301 |
| B2_ALL_lea | LDL_A | Wald ratio | 1 | -0.00811 | 0.030331 | 0.789094 | 0.99301 |
| C2_ALL_lea | Ibil | Wald ratio | 1 | -0.00461 | 0.018058 | 0.798475 | 0.99301 |
| C2_ALL_lea | Tbil | Wald ratio | 1 | -0.00518 | 0.020281 | 0.798475 | 0.99301 |
| B1_ALL_lea | WBC_coun | Wald ratio | 1 | -0.02197 | 0.107644 | 0.838263 | 0.99301 |
| C2_ALL_eu | LDL | Wald ratio | 1 | 0.006455 | 0.032101 | 0.840643 | 0.99301 |
| C2_ALL_eu | LDL_A | Wald ratio | 1 | -0.00407 | 0.020482 | 0.842429 | 0.99301 |
| B1_ALL_lea | APTT | Wald ratio | 1 | 0.009505 | 0.051394 | 0.85327 | 0.99301 |
| B1_ALL_eu | LDL | Wald ratio | 1 | 0.023188 | 0.129346 | 0.857724 | 0.99301 |
| C2_ALL_eu | Ibil | Wald ratio | 1 | 0.002868 | 0.019715 | 0.884338 | 0.99301 |
| C2_ALL_eu | Tbil | Wald ratio | 1 | 0.003221 | 0.022142 | 0.884338 | 0.99301 |
| B2_ALL_lea | Ibil | Wald ratio | 1 | 0.005445 | 0.04384 | 0.901147 | 0.99301 |
| B2_ALL_lea | Tbil | Wald ratio | 1 | 0.006116 | 0.049238 | 0.901147 | 0.99301 |
| C2_ALL_lea | LDL | Wald ratio | 1 | 0.002767 | 0.028449 | 0.922512 | 0.99301 |
| B2_ALL_lea | Ibil | Wald ratio | 1 | -0.0032 | 0.036954 | 0.931023 | 0.99301 |
| B2_ALL_lea | Tbil | Wald ratio | 1 | -0.00359 | 0.041505 | 0.931023 | 0.99301 |
| B1_ALL_lea | Ibil | Wald ratio | 1 | 0.004 | 0.060293 | 0.947103 | 0.99301 |
| B1_ALL_lea | Tbil | Wald ratio | 1 | 0.004493 | 0.067718 | 0.947103 | 0.99301 |
| B1_ALL_eu | APTT | Wald ratio | 1 | 0.003011 | 0.059245 | 0.959462 | 0.99301 |
| B2_ALL_lea | LDL_A | Wald ratio | 1 | 0.000878 | 0.036255 | 0.980675 | 0.99301 |
| C2_ALL_eu | Ibil | Wald ratio | 1 | 0.000207 | 0.023587 | 0.99301 | 0.99301 |
| C2_ALL_eu | Tbil | Wald ratio | 1 | 0.000232 | 0.026492 | 0.99301 | 0.99301 |

S2.BBJ LDL-C one SNP

| outcome | exposure | method | nsnp | b | se | p-value | q-value |
|---|---|---|---|---|---|---|---|
| B2_ALL_eu | LDL-C | Wald ratio | 1 | 0.201381 | 0.072642 | 0.005567 | 0.037598 |
| C2_ALL_lea | LDL-C | Wald ratio | 1 | -0.10377 | 0.037963 | 0.006266 | 0.037598 |
| B2_ALL_lea | LDL-C | Wald ratio | 1 | 0.160644 | 0.066395 | 0.015541 | 0.062164 |
| C2_ALL_eu | LDL-C | Wald ratio | 1 | -0.09088 | 0.041093 | 0.027005 | 0.081016 |
| B1_ALL_eu | LDL-C | Wald ratio | 1 | 0.17349 | 0.113535 | 0.126493 | 0.267541 |
| B2_ALL_lea | LDL-C | Wald ratio | 1 | 0.115547 | 0.081243 | 0.154959 | 0.267541 |
| C2_ALL_lea | LDL-C | Wald ratio | 1 | -0.04398 | 0.031946 | 0.168632 | 0.267541 |
| B1_ALL_lea | LDL-C | Wald ratio | 1 | 0.142175 | 0.105642 | 0.178361 | 0.267541 |
| B2_ALL_eu | LDL-C | Wald ratio | 1 | 0.112893 | 0.092006 | 0.219815 | 0.293087 |
| B1_ALL_lea | LDL-C | Wald ratio | 1 | 0.091025 | 0.15149 | 0.54793 | 0.630338 |
| C2_ALL_eu | LDL-C | Wald ratio | 1 | -0.01893 | 0.034014 | 0.57781 | 0.630338 |
| B1_ALL_eu | LDL-C | Wald ratio | 1 | 0.057496 | 0.170671 | 0.736206 | 0.736206 |

S3.BBJ LDL-C all SNPs

| outcome | exposure | method | nsnp | b | se | p-value | q-value |
|---|---|---|---|---|---|---|---|
| C2_ALL_lea | LDL-C | Inverse var | 11 | -0.086 | 0.033594 | 0.01047 | 0.089814 |
| C2_ALL_eu | LDL-C | Inverse var | 11 | -0.0882 | 0.03625 | 0.014969 | 0.089814 |
| C2_ALL_lea | LDL-C | Inverse var | 12 | -0.03699 | 0.029964 | 0.216962 | 0.734102 |
| B2_ALL_eu | LDL-C | Inverse var | 12 | 0.095809 | 0.087254 | 0.272183 | 0.734102 |
| B1_ALL_eu | LDL-C | Inverse var | 11 | 0.11125 | 0.115132 | 0.333902 | 0.734102 |
| B2_ALL_lea | LDL-C | Inverse var | 12 | 0.064588 | 0.075809 | 0.394219 | 0.734102 |
| C2_ALL_eu | LDL-C | Inverse var | 11 | -0.02605 | 0.032887 | 0.428226 | 0.734102 |
| B1_ALL_lea | LDL-C | Inverse var | 12 | 0.062056 | 0.096383 | 0.519671 | 0.779507 |
| B2_ALL_eu | LDL-C | Inverse var | 12 | -0.01451 | 0.095582 | 0.87938 | 0.980667 |
| B1_ALL_eu | LDL-C | Inverse var | 12 | -0.02163 | 0.148288 | 0.884034 | 0.980667 |
| B1_ALL_lea | LDL-C | Inverse var | 11 | -0.01058 | 0.129443 | 0.934866 | 0.980667 |
| B2_ALL_lea | LDL-C | Inverse var | 12 | -0.00186 | 0.076819 | 0.980667 | 0.980667 |

S4.Cholesterol EUR

| outcome | exposure | method | nsnp | b | se | p-value | q-value |
|---|---|---|---|---|---|---|---|
| B2_ALL_eu | Cholesterol | Wald ratio | 1 | -5.37235 | 2.135392 | 0.011874 | 0.07554 |
| B1_ALL_eu | Cholesterol | Wald ratio | 1 | -8.80765 | 3.634118 | 0.015368 | 0.07554 |
| B2_ALL_eu | Cholesterol | Wald ratio | 1 | -5.85588 | 2.494216 | 0.018885 | 0.07554 |
| B2_ALL_lea | Cholesterol | Wald ratio | 1 | -4.08539 | 1.877451 | 0.029553 | 0.088659 |
| B2_ALL_lea | Cholesterol | Wald ratio | 1 | -4.38284 | 2.132255 | 0.039831 | 0.095594 |
| B1_ALL_lea | Cholesterol | Wald ratio | 1 | -6.34196 | 3.20549 | 0.047876 | 0.095752 |
| B1_ALL_eu | Cholesterol | Wald ratio | 1 | -8.17 | 5.077353 | 0.107593 | 0.184445 |
| B1_ALL_lea | Cholesterol | Wald ratio | 1 | -5.37088 | 4.176863 | 0.19849 | 0.297735 |
| C2_ALL_eu | Cholesterol | Wald ratio | 1 | -1.24235 | 1.315196 | 0.344856 | 0.459808 |
| C2_ALL_eu | Cholesterol | Wald ratio | 1 | -0.79885 | 1.117745 | 0.474794 | 0.569753 |
| C2_ALL_lea | Cholesterol | Wald ratio | 1 | -0.35272 | 1.149608 | 0.758985 | 0.793428 |
| C2_ALL_lea | Cholesterol | Wald ratio | 1 | -0.2622 | 1.001275 | 0.793428 | 0.793428 |

S5.Cholesterol AFR

| outcome | exposure | method | nsnp | b | se | p-value | q-value |
|---|---|---|---|---|---|---|---|
| B1_ALL_lea | Cholestero | Inverse var | 8 | -0.25933 | 0.100015 | 0.009516 | 0.111252 |
| B1_ALL_lea | Cholestero | Inverse var | 8 | -0.17633 | 0.074886 | 0.018542 | 0.111252 |
| B2_ALL_lea | Cholestero | Inverse var | 8 | -0.08224 | 0.045274 | 0.069301 | 0.275174 |
| B2_ALL_eu | Cholestero | Inverse var | 8 | -0.08547 | 0.051844 | 0.099239 | 0.275174 |
| B1_ALL_eu | Cholestero | Inverse var | 8 | -0.1943 | 0.123163 | 0.114656 | 0.275174 |
| B1_ALL_eu | Cholestero | Inverse var | 8 | -0.13884 | 0.094986 | 0.143822 | 0.287644 |
| C2_ALL_eu | Cholestero | Inverse var | 8 | -0.03372 | 0.025812 | 0.191401 | 0.288387 |
| C2_ALL_lea | Cholestero | Inverse var | 8 | -0.03004 | 0.023037 | 0.192258 | 0.288387 |
| B2_ALL_lea | Cholestero | Inverse var | 8 | -0.05054 | 0.048561 | 0.297999 | 0.397332 |
| B2_ALL_eu | Cholestero | Inverse var | 8 | -0.02985 | 0.059471 | 0.615744 | 0.738893 |
| C2_ALL_lea | Cholestero | Inverse var | 8 | -0.00891 | 0.026623 | 0.737914 | 0.743385 |
| C2_ALL_eu | Cholestero | Inverse var | 8 | -0.01005 | 0.030685 | 0.743385 | 0.743385 |

S6.WBC EAS one SNP

| outcome | exposure | method | nsnp | b | se | p-value | q-value |
|---|---|---|---|---|---|---|---|
| C2_ALL_eu | White bloc | Wald ratio | 1 | 0.635212 | 0.178467 | 0.000372 | 0.003144 |
| C2_ALL_lea | White bloc | Wald ratio | 1 | 0.561105 | 0.161786 | 0.000524 | 0.003144 |
| C2_ALL_eu | White bloc | Wald ratio | 1 | 0.612868 | 0.212309 | 0.003893 | 0.015572 |
| C2_ALL_lea | White bloc | Wald ratio | 1 | 0.505148 | 0.18756 | 0.007076 | 0.021228 |
| B2_ALL_eu | White bloc | Wald ratio | 1 | 0.409811 | 0.341259 | 0.229797 | 0.551513 |
| B2_ALL_lea | White bloc | Wald ratio | 1 | 0.300322 | 0.298288 | 0.314021 | 0.601615 |
| B1_ALL_lea | White bloc | Wald ratio | 1 | 0.646941 | 0.693575 | 0.350942 | 0.601615 |
| B1_ALL_lea | White bloc | Wald ratio | 1 | 0.427361 | 0.51992 | 0.411092 | 0.616638 |
| B1_ALL_eu | White bloc | Wald ratio | 1 | 0.507377 | 0.81244 | 0.532293 | 0.641279 |
| B2_ALL_eu | White bloc | Wald ratio | 1 | 0.235502 | 0.402841 | 0.558815 | 0.641279 |
| B1_ALL_eu | White bloc | Wald ratio | 1 | 0.310229 | 0.572409 | 0.587839 | 0.641279 |
| B2_ALL_lea | White bloc | Wald ratio | 1 | 0.118208 | 0.339932 | 0.728036 | 0.728036 |

S7.WBC EAS all SNPs

| outcome | exposure | method | nsnp | b | se | p-value | q-value |
|---|---|---|---|---|---|---|---|
| B2_ALL_lea | White bloc | Inverse var | 48 | 0.198945 | 0.084427 | 0.018452 | 0.18231 |
| B2_ALL_eu | White bloc | Inverse var | 47 | 0.215439 | 0.099508 | 0.030385 | 0.18231 |
| B1_ALL_lea | White bloc | Inverse var | 47 | 0.375939 | 0.194317 | 0.053031 | 0.212124 |
| B1_ALL_lea | White bloc | Inverse var | 47 | 0.252466 | 0.151241 | 0.09506 | 0.236504 |
| C2_ALL_eu | White bloc | Inverse var | 49 | 0.098082 | 0.059515 | 0.099349 | 0.236504 |
| C2_ALL_lea | White bloc | Inverse var | 48 | 0.089337 | 0.057188 | 0.118252 | 0.236504 |
| B2_ALL_lea | White bloc | Inverse var | 46 | 0.15621 | 0.105434 | 0.13845 | 0.237343 |
| B2_ALL_eu | White bloc | Inverse var | 47 | 0.171592 | 0.131871 | 0.193187 | 0.289781 |
| B1_ALL_eu | White bloc | Inverse var | 48 | 0.183794 | 0.17314 | 0.288447 | 0.373577 |
| B1_ALL_eu | White bloc | Inverse var | 48 | 0.253371 | 0.250251 | 0.311314 | 0.373577 |
| C2_ALL_eu | White bloc | Inverse var | 49 | 0.062835 | 0.068625 | 0.359865 | 0.39258 |
| C2_ALL_lea | White bloc | Inverse var | 48 | 0.051625 | 0.063534 | 0.416468 | 0.416468 |

S8.Reverse MR with LDL-C

| outcome | exposure | method | nsnp | b | se | p-value |
|---|---|---|---|---|---|---|
| LDL | B2_ALL_eu | Inverse var | 2 | -0.32678 | 0.468397 | 0.485398 |
| LDL | B2_ALL_eu | Inverse var | 4 | -0.0856 | 0.313749 | 0.784979 |
| LDL | B2_ALL_lea | Inverse var | 6 | 0.089936 | 0.36142 | 0.803484 |
| LDL | B2_ALL_lea | Inverse var | 3 | -0.20535 | 0.371473 | 0.580393 |
| LDL | C2_ALL_eu | Inverse var | 4 | -0.28729 | 0.617801 | 0.641918 |
| LDL | C2_ALL_eu | Inverse var | 2 | -0.63887 | 0.877206 | 0.466432 |
| LDL | C2_ALL_lea | Inverse var | 6 | -0.18688 | 0.63396 | 0.768163 |
| LDL | C2_ALL_lea | Inverse var | 2 | -0.77048 | 1.005556 | 0.443545 |
| LDL | B1_ALL_eu | Wald ratio | 1 | -0.34853 | 0.589781 | 0.554557 |

S9.Reverse MR with WBC count

| outcome | exposure | method | nsnp | b | se | p-value |
|---|---|---|---|---|---|---|
| WBC | B2_ALL_eu | Inverse var | 2 | -0.12327 | 0.896405 | 0.890625 |
| WBC | B2_ALL_eu | Inverse var | 4 | 0.071301 | 0.378142 | 0.85044 |
| WBC | B2_ALL_lea | Inverse var | 6 | 0.204908 | 0.218167 | 0.347616 |
| WBC | B2_ALL_lea | Inverse var | 3 | 0.17189 | 0.533219 | 0.747177 |
| WBC | C2_ALL_eu | Inverse var | 4 | 0.381635 | 0.610495 | 0.53189 |
| WBC | C2_ALL_eu | Inverse var | 2 | -0.36783 | 1.539663 | 0.811179 |
| WBC | C2_ALL_lea | Inverse var | 6 | 0.457057 | 0.625305 | 0.464819 |
| WBC | C2_ALL_lea | Inverse var | 2 | -0.3302 | 1.785542 | 0.853284 |
| WBC | B1_ALL_eu | Wald ratio | 1 | 1.110708 | 0.585999 | 0.058038 |
