## Supplementary figures and images for "A Chinese host genetic study discovered type I interferons and causality of cholesterol levels and WBC counts on COVID-19 severity"

### supplementary figures.pdf

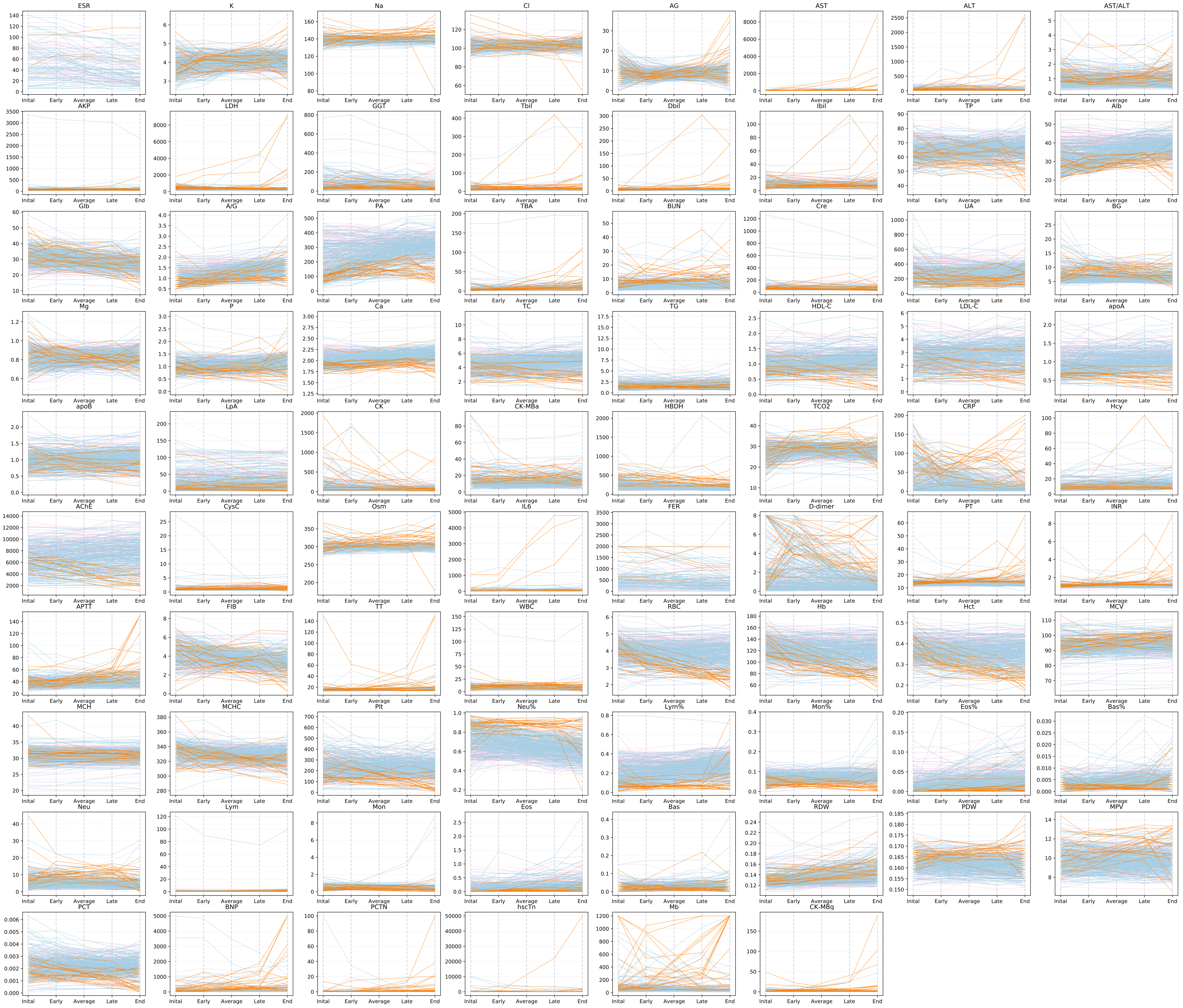

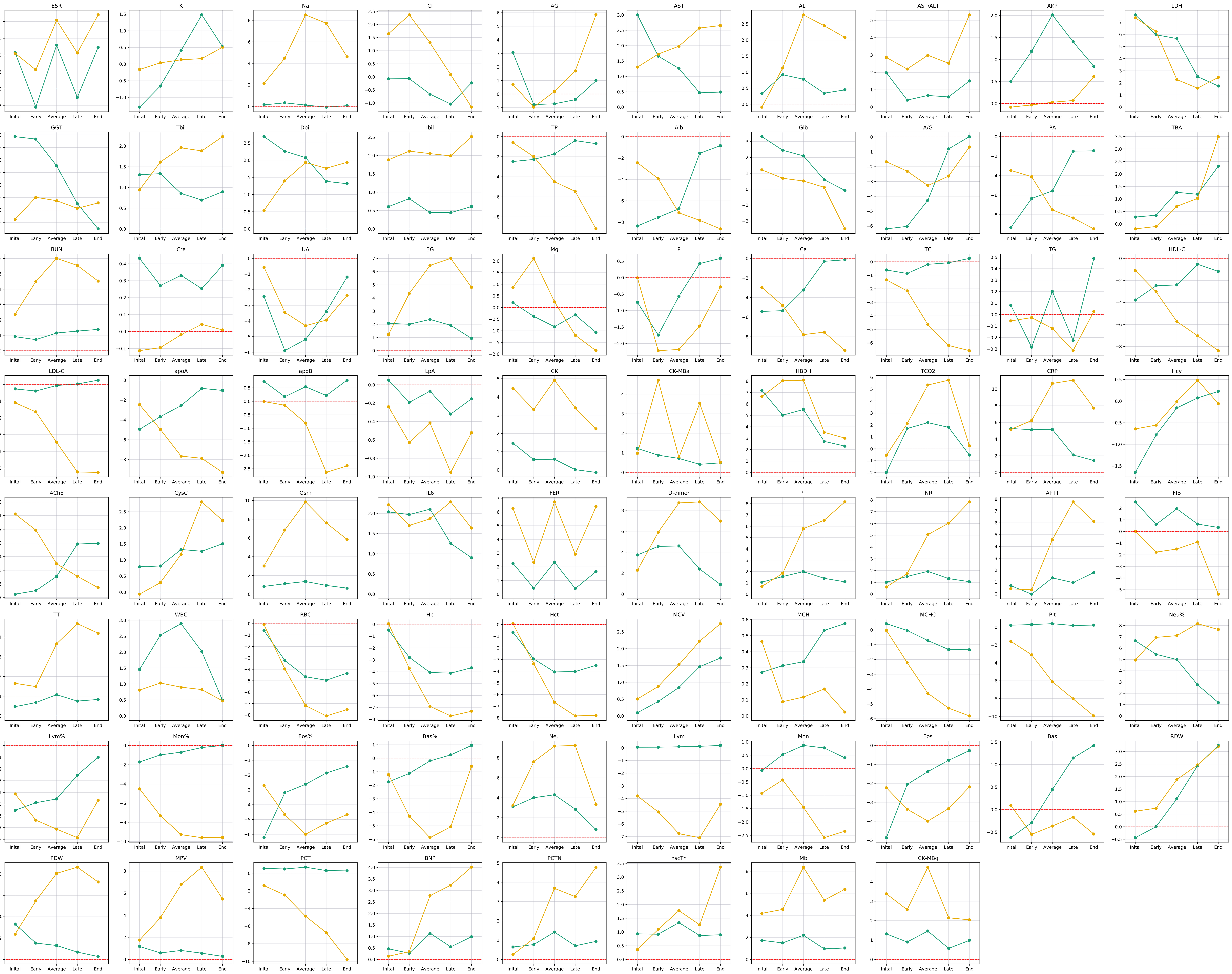
